# Effect of common pregnancy and perinatal complications on offspring metabolic traits across the life course: a multi-cohort study

**DOI:** 10.1101/2022.04.05.22273388

**Authors:** Ahmed Elhakeem, Justiina Ronkainen, Toby Mansell, Katherine Lange, Tuija Mikkola, Binisha H Mishra, Rama Wahab, Tim Cadman, Tiffany Yang, David Burgner, Johan G Eriksson, Marjo-Riitta Järvelin, Romy Gaillard, Vincent WV Jaddoe, Terho Lehtimäki, Olli T Raitakari, Richard Saffery, Melissa Wake, John Wright, Sylvain Sebert, Deborah A Lawlor

## Abstract

**Backgroun:** Common pregnancy and perinatal complications are associated with offspring cardiometabolic risk factors. These complications may influence multiple metabolic traits in offspring and these associations might differ with offspring age.

**Methods:** We used data from eight population-based cohort studies to examine and compare associations of pre-eclampsia (PE), gestational hypertension (GH), gestational diabetes (GD), preterm birth (PTB), small (SGA) and large (LGA) for gestational age (vs. appropriate size for gestational age (AGA)) with up to 167 plasma/serum-based nuclear magnetic resonance-derived metabolic traits encompassing lipids, lipoproteins, fatty acids, amino acids, ketones, glycerides/phospholipids, glycolysis, fluid balance, and inflammation. Confounder-adjusted regression models were used to examine associations (adjusted for maternal education, parity age at pregnancy, ethnicity, pre/early pregnancy body mass index and smoking, and offspring sex and age at metabolic trait assessment), and results were combined using meta-analysis by five age categories representing different periods of the offspring life course: neonates (cord blood), infancy (mean ages: 1.1-1.6y), childhood (4.2-7.5y); adolescence (12.0-16.0y), and adulthood (22.0-67.8y).

**Results:** Offspring numbers for each age category/analysis varied from 8,925 adults (441 PTB) to 1,181 infants (135 GD); 48.4% to 60.0% were females. Pregnancy complications (PE, GH, GD) were each associated with up to three metabolic traits in neonates (*P*≤0.001) with some limited evidence of persistence to older ages. PTB and SGA were associated with 32 and 12 metabolic traits in neonates respectively, which included an adjusted standardised mean difference of -0.89 standard deviation (SD) units for *albumin* with PTB (95%CI: -1.10 to -0.69, *P*=1.3×10^−17^) and -0.41SD for *total lipids in medium HDL* with SGA (95%CI: -0.56 to -0.25, *P*=2.6×10^−7^), with limited evidence of persistence to older ages. LGA was inversely associated with 19 metabolic traits including lower levels of cholesterol, lipoproteins, fatty acids, and amino acids, with associations emerging in adolescence, (e.g., -0.11SD *total fatty acids*, 95%CI: -0.18 to -0.05, *P*=0.0009), and attenuating with older age across adulthood.

**Conclusions:** These reassuring findings suggest little evidence of wide-spread and long-term impact of common pregnancy and perinatal complications on offspring metabolic traits, with most associations only observed for new-borns rather than older ages, and for perinatal rather than pregnancy complications.

## INTRODUCTION

There are widespread changes in maternal circulating metabolites during pregnancy, which return to normal after pregnancy (1). These alterations are likely to be important for maternal health, normal fetal growth, and development (2). Maternal metabolic profiles are associated with different common pregnancy and perinatal complications including pre-eclampsia (PE), gestational hypertension (GH), gestational diabetes (GD), preterm birth (PTB) and small (SGA) and large for gestational age (LGA) (3-9). PE, GH, PTB, and SGA tend to relate to placental pathologies/fetal growth restriction (10, 11), whereas GD and LGA relate to fetal overgrowth (12, 13). Both fetal growth restriction and overgrowth might have long-lasting metabolic effects, which may in turn increase cardiovascular disease (CVD) risk (14-16).

Studies indicate common pregnancy/perinatal complications associate with cardiovascular disease in the offspring (17-22). However, to the best of our knowledge, effects of common pregnancy/perinatal complications on offspring metabolic traits, and whether these change with age, have not been examined. Identifying whether effects on offspring metabolism are short lived, persist across life, emerge later, or strengthen/weaken with age can improve our understanding of CVD aetiology and may inform timing of interventions. Therefore, the aim of this study was to examine associations of common pregnancy and perinatal complications related to fetal growth restriction (PE, GH, PTB, and SGA) and fetal overgrowth (GD, LGA) with targeted metabolomic profiles across the offspring life course and investigate whether associations differ by offspring age at metabolite assessment.

## METHODS

This study was carried out by following a pre-specified analysis plan and code developed by AE and DAL (https://osf.io/vfd7g) and is reported in accordance with The Strengthening the Reporting of Observational Studies in Epidemiology (STROBE) Statement guidelines for cohort studies (23).

### Cohort studies

Participating cohorts were recruited from the EU Child Cohort Network (EUCCN) (24); a consortium of European and Australian pregnancy/birth cohorts. Studies were included if they had data on (i) at least one pregnancy/perinatal complication, (ii) offspring metabolic profiles measured at any age in plasma/serum by high-throughput proton nuclear magnetic resonance (NMR)-based targeted metabolomics platform (most widely used platform across EUCCN cohorts) (25), and (iii) prespecified confounders. Eight cohorts were eligible and all agreed to participate in this analysis: the UK-based Avon Longitudinal Study of Parents and Children (ALSPAC) (26-30), and Born in Bradford Study (BiB) (31, 32), the Finland-based Young Finns Study (YFS) (33-35), Northern Finland Birth Cohort 1966 (NFBC1966) (35-37), Northern Finland Birth Cohort 1986 (NFBC1986) (37), and the Helsinki Birth Cohort Study (HBCS) (38), and the Australia-based Barwon Infant Study (BIS) (36, 37), and the Longitudinal Study of Australian Children’s Child Health CheckPoint (CheckPoint) (39, 40).

All cohorts had ethical approval from their relevant local or national ethics committees and study participants provided informed consent or assent to participate in the respective cohorts and secondary data analyses. Cohort descriptions and details on ethics approvals and consent for each cohort can be found in the **Supplemental Methods**.

### Pregnancy and perinatal complications

Six common pregnancy/perinatal complications related to fetal growth restriction (PE, GH, PTB, and SGA) and fetal overgrowth (GD, LGA) were included. Description of how these were measured in each cohort is in the **Supplemental Methods**. Data harmonisation was previously described (41). Briefly, PE was defined as elevated blood pressure >20 weeks gestation (≥140 mmHg systolic or ≥90 mmHg diastolic), and proteinuria (>0.3g per 24 h), or by HELLP syndrome (haemolysis, elevated liver enzymes, and low platelets) (42). GH was defined as new onset hypertension >20 weeks of gestation, with previously normal blood pressure, without proteinuria or manifestations of PE. GD was defined as glucose intolerance with onset or first diagnosis in pregnancy and continuing beyond 24–28 weeks of gestation. PTB was calculated as ≤37 completed weeks (or ≤259 days) at birth. SGA and LGA were defined based on the World Health Organisation fetal growth charts, using the 5^th^ and 95^th^ percentiles as cut-offs, respectively (43).

For PE, GH, PTB, and GD, four variables were derived to compare offspring exposed to each complication with non-exposed offspring. For SGA and LGA, two variables were derived to compare those born SGA or LGA with appropriate size for gestational age (AGA) offspring i.e., with SGA/LGA excluded in turn.

### Offspring NMR-derived metabolic traits and age categories

A proton NMR-based targeted metabolomics platform (25) was used to quantify up to 250 offspring metabolic traits (including derived variables) in plasma/serum samples in the eight participating cohorts. Metabolic traits were quantified in absolute concentration units or ratios and included circulating lipoprotein lipids and subclasses, fatty acids and their compositions, amino acids and traits related to glycolysis, ketone bodies, fluid balance, and an inflammatory marker. Traits were analysed using non-fasting samples in infancy, semi-fasting samples in childhood and adolescence, and fasting samples at older ages. The manufacturer’s standard quality control procedures were performed in all cohorts (25). Metabolic trait ratios were excluded because of challenges in their interpretation, leaving up to 167 metabolic traits in the analysis. Description of the methods and ages at assessment of metabolic traits in each cohort is in **Supplemental Methods**. A list of the metabolic traits available within each cohort is provided in **Data Set 1**.

Metabolic traits measured at all available ages were included. Results were combined into five pre-specified age categories chosen to reflect key life course periods and to maximise numbers of participants **(Table 2)**. The age categories were neonates (cord blood), infancy (mean age 1.1 to 1.6 years), childhood (mean age 4.2 to 7.5 years), adolescence (mean age 12.0 to 16.0 years), and adulthood (mean age 22.0 to 67.8 years).

### Confounders

To estimate unconfounded associations of pregnancy/perinatal complications with offspring metabolic traits, we identified and adjusted for potential confounders i.e., factors that could plausibly cause pregnancy/perinatal complications and influence offspring metabolism, and avoided adjustment for mediators on causal path of any effect (e.g., offspring adiposity) and other sources of collider bias (44). The identified confounders were maternal education (the most available and consistent indicator of early life socioeconomic position across cohorts), ethnicity, age at pregnancy/birth of offspring, parity, pre/early pregnancy body mass index (BMI) and smoking in pregnancy. Offspring sex and age at metabolic trait assessment were included as adjustments to improve modelling precision. Details on how confounders were measured in each cohort is provided in **Supplemental Methods**. Harmonised variables were derived as described previously (41). ALSPAC, BiB, BIS, and YFS were able to adjust for all confounders. NFBC1966, NFBC1986, and HBCS did not adjust for ethnicity, but they were predominantly white ethnicity, HBCS was unable to adjust for smoking, and CheckPoint was unable to adjust for BMI or parity.

### Statistical analysis

Associations between each of the six pregnancy/perinatal complications and each offspring NMR-derived metabolic trait from all available timepoints were examined in each cohort by fitting adjusted (for confounders plus offspring age and sex) linear regression models (with robust standard errors). Analyses were restricted to those with complete data on the relevant pregnancy/perinatal complication, metabolic trait, and confounders. To allow comparison of results across different pregnancy/perinatal complications, traits, and ages, metabolic traits were analysed in cohort-specific standard deviation (SD) units (mean=0, SD=1).

Cohort-specific results were then combined using meta-analysis in five age categories for neonates, infancy, childhood, adolescence, and adulthood, using a random effects model to allow for between-cohort heterogeneity. Variability in the meta-analysis results that was due to between-cohort heterogeneity was measured by computing the *I*^*2*^ statistic (45, 46). Where evidence of substantial between-cohort heterogeneity was found, we inspected each cohort’s results to identify reason for heterogeneity. A *P*-value threshold of *P*≤0.001 was selected to identify statistically robust associations between pregnancy/perinatal complications and metabolic traits. This was chosen instead of a Bonferroni-corrected *P*-value because many metabolic traits are highly correlated (40) and so independent tests were not performed, yet our threshold is still more stringent than a conventional *P*-value threshold. For association that reached this threshold in one age category, we highlighted the equivalent association in all other age categories, to explore changes with age in the context of different numbers of participants for each age category (**Table 1**).

**Table 1.** Characteristics of the cohorts and offspring included in the analysis*

|  |  |  |  |  |  |  |  |  |
| --- | --- | --- | --- | --- | --- | --- | --- | --- |
| *Cohort name* | ALSPAC | BiB | YFS | NFBC1986 | NFBC1966 | HBCS | BIS | CheckPoint |
| *Cohort country* | UK | UK | Finland | Finland | Finland | Finland | Australia | Australia |
| *Offspring birth years* | 1990-1992 | 2007-2011 | 1962-1977 | 1985-1986 | 1966 | 1934-1944 | 2010-2013 | 2003-2004 |
| *Mean age(s) (in years) at assessment of NMR-derived metabolic traits* | 7.5y, 15.4y,<br>17.8y, 24.5y | cord blood,<br>1.6y | 22.0y | 16.0y | 31.2y, 46.6y | 67.8y | cord blood,<br>1.1y, 4.2y | 12.0y |
| *Sex [No. (%)]* |  |  |  |  |  |  |  |  |
| male | 3216 (49.6) | 1290 (51.6) | 204 (40.0%) | 2305 (50.0) | 2400 (47.9) | 466 (44.0%) | 374 (51.5) | 429 (47.8) |
| female | 3263 (50.4) | 1209 (48.4) | 305 (60.0%) | 2309 (50.0) | 2607 (52.1) | 593 (56.0%) | 352 (48.5) | 469 (52.2) |
| *Pre-eclampsia [No. (%)]* |  |  |  |  |  |  |  |  |
| no | 6335 (98.1) | 2328 (97.2) | 499 (98.0%) | 3596 (97.5) | 2883 (96.3) | - | 701 (97.0) | - |
| yes | 120 (1.9) | 66 (2.8) | 10 (2.0%) | 91 (2.5) | 110 (3.7) | - | 22 (3.0) | - |
| *Gestational hypertension [No. (%)]* |  |  |  |  |  |  |  |  |
| no | 5433 (85.8) | 2215 (92.5) | 492 (97.0%) | 3596 (96.4) | 2883 (86.9) | - | 713 (98.2) | 843 (94.2) |
| yes | 902 (14.2) | 179 (7.5) | 17 (3.0%) | 134 (3.6) | 406 (13.1) | - | 13 (1.8) | 52 (5.8) |
| *Gestational diabetes [No. (%)]* |  |  |  |  |  |  |  |  |
| no | 6424 (99.5) | 1091 (84.7) | 468 (92%) | - | - | - | 582 (95.7) | 846 (95.1) |
| yes | 31 (0.5) | 197 (15.3) | 41 (8%) | - | - | - | 26 (4.3) | 44 (4.9) |
| *Preterm birth [No. (%)]* |  |  |  |  |  |  |  |  |
| no | 6156 (95.0) | 2342 (93.7) | 460 (90.0%) | 4381 (95.0) | 4775 (95.4) | 1000 (94.4) | 682 (93.9) | 800 (89.6) |
| yes | 323 (5.0) | 157 (6.3) | 49 (10.0%) | 230 (5.0) | 225 (4.6) | 59 (5.6) | 44 (6.0) | 93 (10.4) |
| *Small for gestational age [No. (%)]* |  |  |  |  |  |  |  |  |
| no (appropriate size for age) | 5446 (94.4) | 2140 (90.8) | - | 3842 (96.5) | 3928 (94.5) | - | 631 (98.3) | 764 (95.3) |
| yes | 323 (5.6) | 216 (9.2) | - | 138 (3.5) | 227 (5.6) | - | 11 (1.7) | 38 (4.7) |
| *Large for gestational age [No. (%)]* |  |  |  |  |  |  |  |  |
| no (appropriate size for age) | 5446 (89.9) | 2140 (93.8) | - | 3842 (86.1) | 3928 (89.6) | - | 631 (88.3) | 764 (89.6) |
| yes | 610 (10.1) | 142 (6.2) | - | 621 (13.9) | 454 (10.4) | - | 84 (11.8) | 89 (10.4) |

\* Characteristics presented for study participants with data on at least 1 pregnancy/perinatal complication, NMR-derived metabolic traits, and all available confounders. For the cohort studies that had metabolic traits measured at multiple timepoints, the table shows the characteristics for the study participants with data from the timepoint with the largest sample size.

**Table 2.**
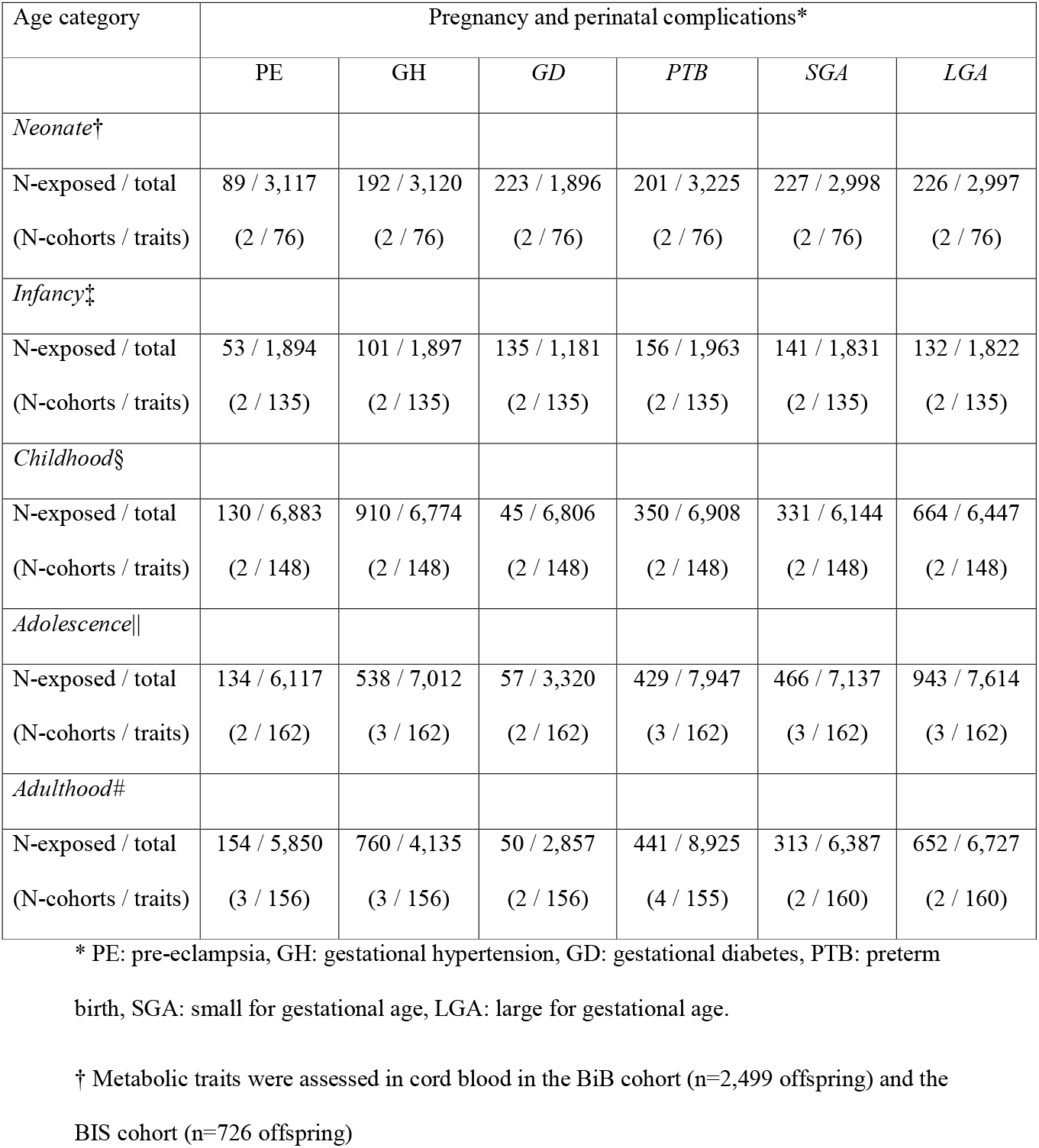
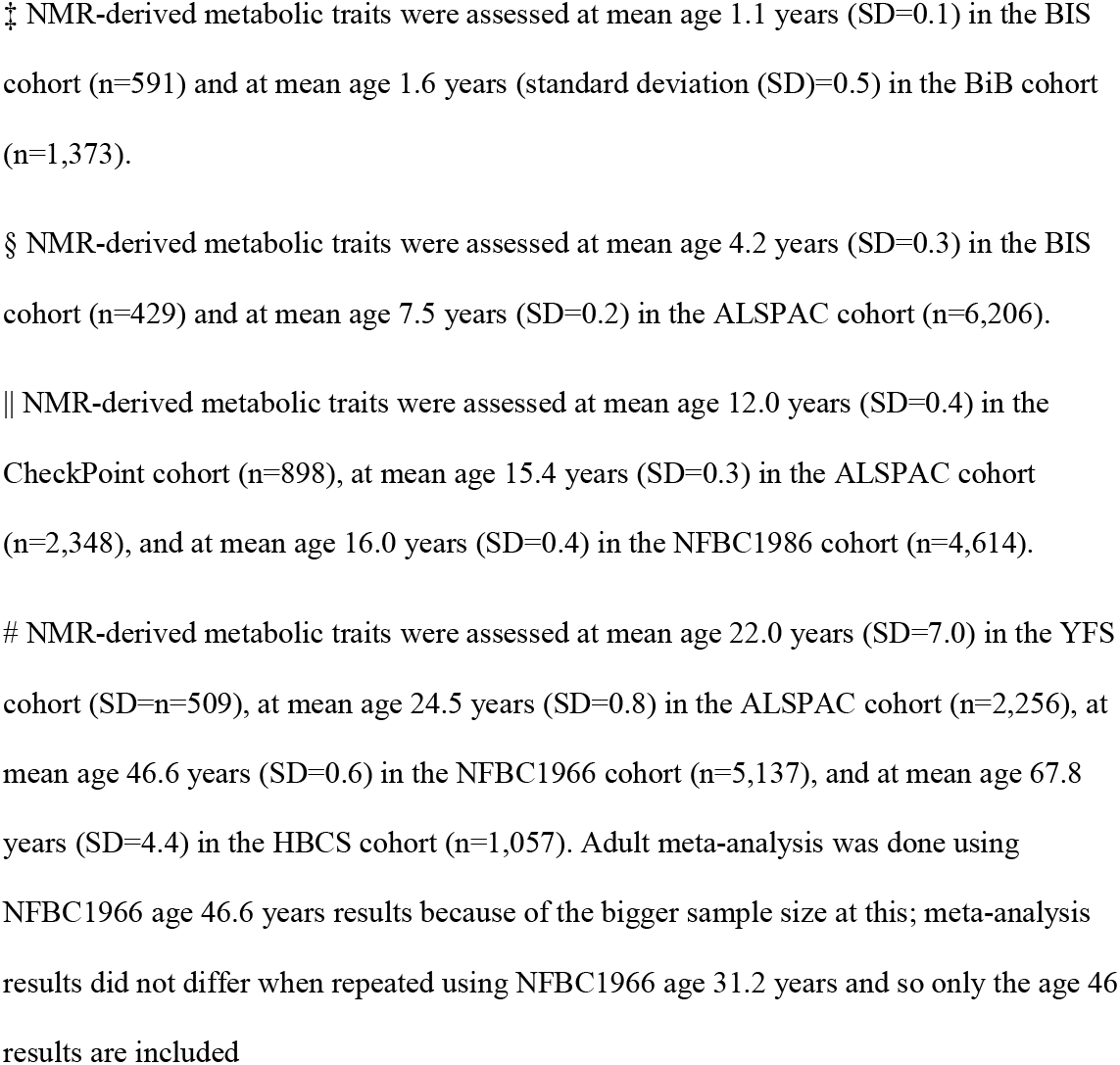
Number of cohorts, offspring, and NMR-derived metabolic traits included in each life course stage analysis

| Age category | Pregnancy and perinatal complications* |  |  |  |  |  |
| --- | --- | --- | --- | --- | --- | --- |
|  | PE | GH | GD | PTB | SGA | LGA |
| *Neonate*<sup>†</sup> |  |  |  |  |  |  |
| N-exposed / total | 89 / 3,117 | 192 / 3,120 | 223 / 1,896 | 201 / 3,225 | 227 / 2,998 | 226 / 2,997 |
| (N-cohorts / traits) | (2 / 76) | (2 / 76) | (2 / 76) | (2 / 76) | (2 / 76) | (2 / 76) |
| *Infancy*<sup>‡</sup> |  |  |  |  |  |  |
| N-exposed / total | 53 / 1,894 | 101 / 1,897 | 135 / 1,181 | 156 / 1,963 | 141 / 1,831 | 132 / 1,822 |
| (N-cohorts / traits) | (2 / 135) | (2 / 135) | (2 / 135) | (2 / 135) | (2 / 135) | (2 / 135) |
| *Childhood*<sup>§</sup> |  |  |  |  |  |  |
| N-exposed / total | 130 / 6,883 | 910 / 6,774 | 45 / 6,806 | 350 / 6,908 | 331 / 6,144 | 664 / 6,447 |
| (N-cohorts / traits) | (2 / 148) | (2 / 148) | (2 / 148) | (2 / 148) | (2 / 148) | (2 / 148) |
| *Adolescence* |  |  |  |  |  |  |
| N-exposed / total | 134 / 6,117 | 538 / 7,012 | 57 / 3,320 | 429 / 7,947 | 466 / 7,137 | 943 / 7,614 |
| (N-cohorts / traits) | (2 / 162) | (3 / 162) | (2 / 162) | (3 / 162) | (3 / 162) | (3 / 162) |
| *Adulthood*<sup>#</sup> |  |  |  |  |  |  |
| N-exposed / total | 154 / 5,850 | 760 / 4,135 | 50 / 2,857 | 441 / 8,925 | 313 / 6,387 | 652 / 6,727 |
| (N-cohorts / traits) | (3 / 156) | (3 / 156) | (2 / 156) | (4 / 155) | (2 / 160) | (2 / 160) |

\* PE: pre-eclampsia, GH: gestational hypertension, GD: gestational diabetes, PTB: preterm
birth, SGA: small for gestational age, LGA: large for gestational age.
<sup>†</sup> Metabolic traits were assessed in cord blood in the BiB cohort (n=2,499 offspring) and the
BIS cohort (n=726 offspring)
‡ NMR-derived metabolic traits were assessed at mean age 1.1 years (SD=0.1) in the BIS cohort (n=591) and at mean age 1.6 years (standard deviation (SD)=0.5) in the BiB cohort (n=1,373).
§ NMR-derived metabolic traits were assessed at mean age 4.2 years (SD=0.3) in the BIS cohort (n=429) and at mean age 7.5 years (SD=0.2) in the ALSPAC cohort (n=6,206).
|| NMR-derived metabolic traits were assessed at mean age 12.0 years (SD=0.4) in the CheckPoint cohort (n=898), at mean age 15.4 years (SD=0.3) in the ALSPAC cohort (n=2,348), and at mean age 16.0 years (SD=0.4) in the NFBC1986 cohort (n=4,614).

We further investigated change in associations with older age by fitting confounder-adjusted natural cubic spline mixed effects trajectory models (47) in 4,980 ALSPAC offspring with up to 4 repeated NMR-based assessments from 7 to 26 years. Because ALSPAC data spans ages 7-26 years, trajectory analysis was only done for the metabolic traits showing a meta-analysis association in childhood, adolescence, or adulthood (not neonates or infancy). An interaction between the pregnancy/perinatal complication and age was used to allow different trajectories for exposed/nonexposed offspring. Mean predicted trajectories and differences were obtained from these models (48).

Lastly, we sought support for associations identified with NMR-derived traits by performing a replication analysis in an independent cohort – the Dutch Generation R study – where traits were measured by mass spectrometry in cord blood and in non-fasting blood samples at mean age 9.8 years (48-50). Further details on the Generation R study including mass spectrometer measurements are in the **Supplemental Methods**. Replication was done for all NMR-derived traits that were available from the mass spectrometry platform, using regression models fitted with similar adjustments for confounders.

## RESULTS

The proportion of cohort offspring exposed to each pregnancy/perinatal complication ranged from 1.9% to 3.7% PE, 1.5% to 14.2% GH, 0.5% to 15.3% GD, 4.6% to 10.4% PTB, 3.5% to 9.2% SGA, and 6.2% to 13.9% LGA (**Table 1)**. Offspring birth years were between 1934 and 2013, and 48.4% to 60.0% were female (**Table 1)**. Offspring numbers in each age category analysis varied from 8,925 adults (441 PTB) to 1,181 infants (135 GD) (**Table 2**).

### Pre-eclampsia, gestational hypertension, preterm birth and small for gestational age associations with offspring metabolic traits

PE, GH, PTB, and SGA were associated with 1, 1, 32, and 12 metabolic traits, respectively, with all but 3 of these associations observed for neonates only. PE was associated with lower levels of aromatic amino acid *phenylalanine* in infancy (mean difference: -0.44SD, 95%CI: - 0.67 to -0.22, *P*=0.0002), with differences close to zero in neonates, children, and adults, but there was some evidence in favour of lower levels in adolescents (mean difference: -0.17SD, 95%CI: -0.34 to -0.01, *P*=0.03) (**Figure 1, Table S1**). GH was inversely associated with the ketone *acetate* in infants (mean difference: -0.43SD, 95%CI: -0.59 to -0.28, *P*=3.7×10^−8^), although equivalent differences for other age groups were either close to zero or were imprecisely estimated (**Figure 1, Table S1**).

**Figure 1.**
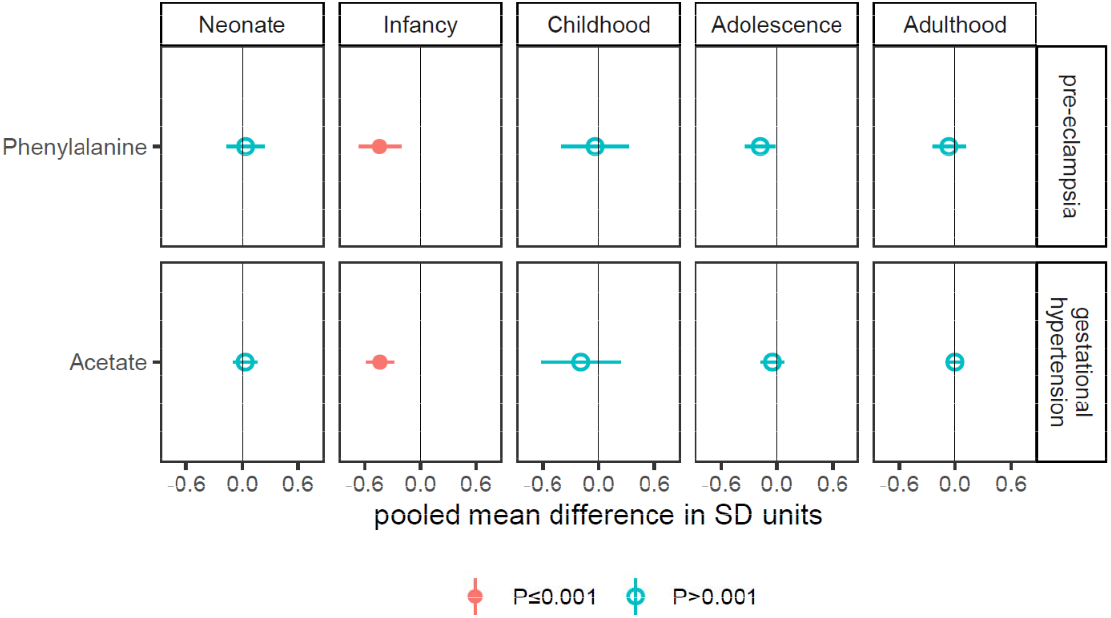
Associations of pre-eclampsia and gestational hypertension with offspring NMR-derived metabolic traits* * Figure shows the pooled adjusted mean differences in standard deviation (SD) units in offspring NMR-derived metabolic traits for pre-eclampsia (minus no pre-eclampsia) and gestational hypertension (minus no gestational hypertension), for associations reaching the *P*<0.001 threshold in any one of the five age categories, and equivalent associations in all other age categories (to explore differences by age). Results are adjusted for offspring sex age, and confounders (maternal education, parity age at pregnancy, ethnicity, pre/early pregnancy BMI and smoking). Horizontal bars represent the 95% confidence intervals. Numerical values of these differences are presented in **Table S1**.

PTB was inversely associated with *total lipids in small HDL, concentration of small HDL particles, degree of unsaturation, glucose*, fluid balance markers *creatinine* and *albumin*, and inflammatory marker *glycoprotein acetyls* (*GlycA*), and positively associated with cholesterol measures, lipoprotein subclasses, glycerides, phospholipids, *Apolipoprotein B, saturated fatty acids*, the α-amino acid *glutamine*, and the aromatic amino acid *tyrosine*, with all associations observed in neonates (**Figure 2**). Associations with neonate metabolic traits for PTB (vs. not PTB) ranged in magnitude from 0.22SD (95%CI: 0.09 to 0.35, *P*=0. 0007) for *total cholines* to -0.89SD (95%CI: -1.10 to -0.69, *P*=1.3×10^−17^) for *albumin*. For most results, equivalent differences across older age categories were close to zero though there was some evidence of persistence to older ages for some metabolic traits, e.g., the mean difference in *total lipids in small LDL* in neonates and adults was 0.33SD (95%CI: 0.16 to 0.50, *P*=0.0001), and 0.14SD (95%CI: 0.04 to 0.24, *P*=0.004), respectively (**Figure 2, Table S1**).

**Figure 2.**
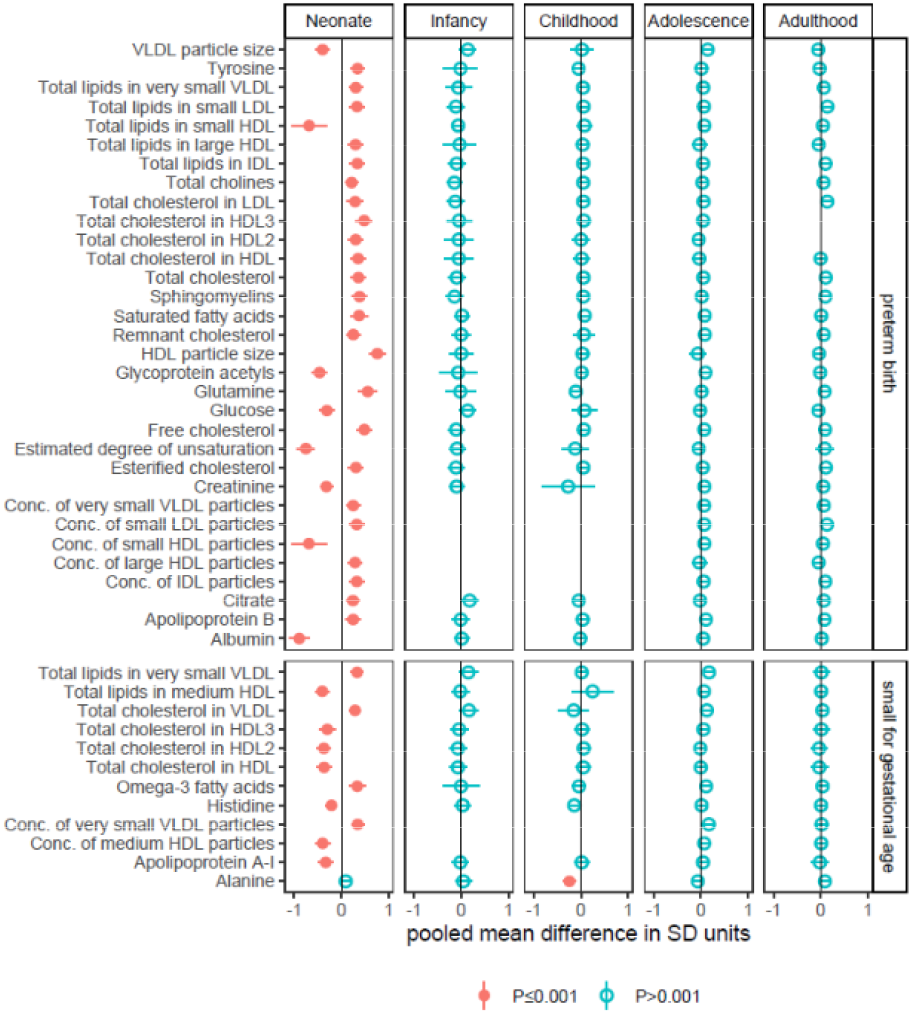
Associations of preterm birth and small for gestational age with offspring NMR-derived metabolic traits* * Figure shows the pooled adjusted mean differences in standard deviation (SD) units in offspring NMR-derived metabolic traits for preterm birth (minus not preterm birth), and small for gestational age (minus appropriate size for gestational age), for associations reaching the *P*<0.001 threshold in any one of the five age categories, and equivalent associations in all other age categories (to explore differences by age). Results are adjusted for offspring sex age, and confounders (maternal education, parity age at pregnancy, ethnicity, pre/early pregnancy BMI and smoking). Horizontal bars represent the 95% confidence intervals. Numerical values of these differences are presented in **Table S1**.

SGA (vs. AGA) in neonates was inversely associated *with total cholesterol in HDL, total cholesterol in HDL2, total cholesterol in HDL3, total lipids in medium HDL, concentration of medium HDL particles, apolipoprotein A-I*, and *histidine*, and positively associated with *total cholesterol in VLDL, total lipids in very small VLDL, concentration of very small VLDL particles*, and *omega-3 fatty acids* (**Figure 2**). Differences ranged in magnitude from -0.21SD (95%CI: -0.33 to -0.08, *P*=0.001) for *histidine* to -0.41SD (95%CI: -0.56 to -0.25, *P*=2.6×10^−7^) for *total lipids in medium HDL* (**Table S1**). Most were reduced at older ages but there was some evidence for higher levels of *total cholesterol in VLDL, total lipids in very small VLDL, concentration of very small VLDL particles*, and *omega-3 fatty acids* during adolescence e.g., mean differences in *total lipids in very small VLDL* in neonates and adolescence were 0.34SD (95%CI: 0.20 to 0.47, *P*=1.4×10^−6^), and 0.17SD (95%CI: 0.06 to 0.29, *P*=0.003), respectively. SGA was also inversely associated with amino acid *alanine* in childhood (−0.25SD, 95%CI: - 0.38 to -0.11, *P*=0.0003), with no clear differences in *alanine* at other age groups (**Figure 2**).

Of the NMR-derived metabolic traits that PE, GH, PTB, and SGA were associated with, four were found among mass spectroscopy measures in the Generation R Study and included for replication (PTB: *tyrosine, sphingomyelins, glutamine*; SGA: *histidine*). Consistent with the pooled difference for PTB neonates in NMR-derived *tyrosine* (0.34SD, 95%CI: 0.19 to 0.49, *P*=7.0×10^−6^) and *sphingomyelins* (0.39SD, 95%CI: 0.23 to 0.54, *P*=6.8×10^−7^), PTB was also associated with higher *tyrosine* (0.82SD, 95%CI: 0.40 to 1.24, *P*=0.0001, n=725 (29 PTB)) and *sphingomyelins* (0.49SD, 95%CI: 0.07 to 0.90, *P*=0.02) in neonates in Generation R. In contrast, the meta-analysis association of PTB with *glutamine* (0.56SD, 95%CI: 0.36 to 0.76, *P*=2.4×10^−8^) did not replicate in Generation R (−0.09, 95%CI: -0.51 to 0.34, *P*=0.7). Lastly, mean difference in *histidine* for SGA (vs. AGA) neonates was similar in the meta-analysis (−0.21SD, 95%CI: -0.33 to -0.08, *P*=0.001) and Generation R but this result was imprecisely estimated (−0.25SD, 95%CI: -0.61 to 0.11, *P*=0.2, n=651 (35 SGA)).

### Gestational diabetes and large for gestational age associations with offspring metabolic traits

GD and LGA were associated with 3 and 19 metabolic traits, respectively. GD was associated with smaller *LDL particle size* (mean difference: -0.25SD, 95%CI: -0.39 to -0.10, *P*=0.0007) and with lower *isoleucine* (mean difference: -0.27SD, 95%CI: -0.41 to -0.14, *P*=0.00008) in neonates, with differences in both metabolic traits close to zero for older ages (**Figure 3**). GD was positively associated with *glucose* in infants (mean difference: 0.35SD, 95%CI: 0.18 to 0.52, *P*=0.00005), with no difference in *glucose* found for other age categories (**Figure 3**).

**Figure 3.**
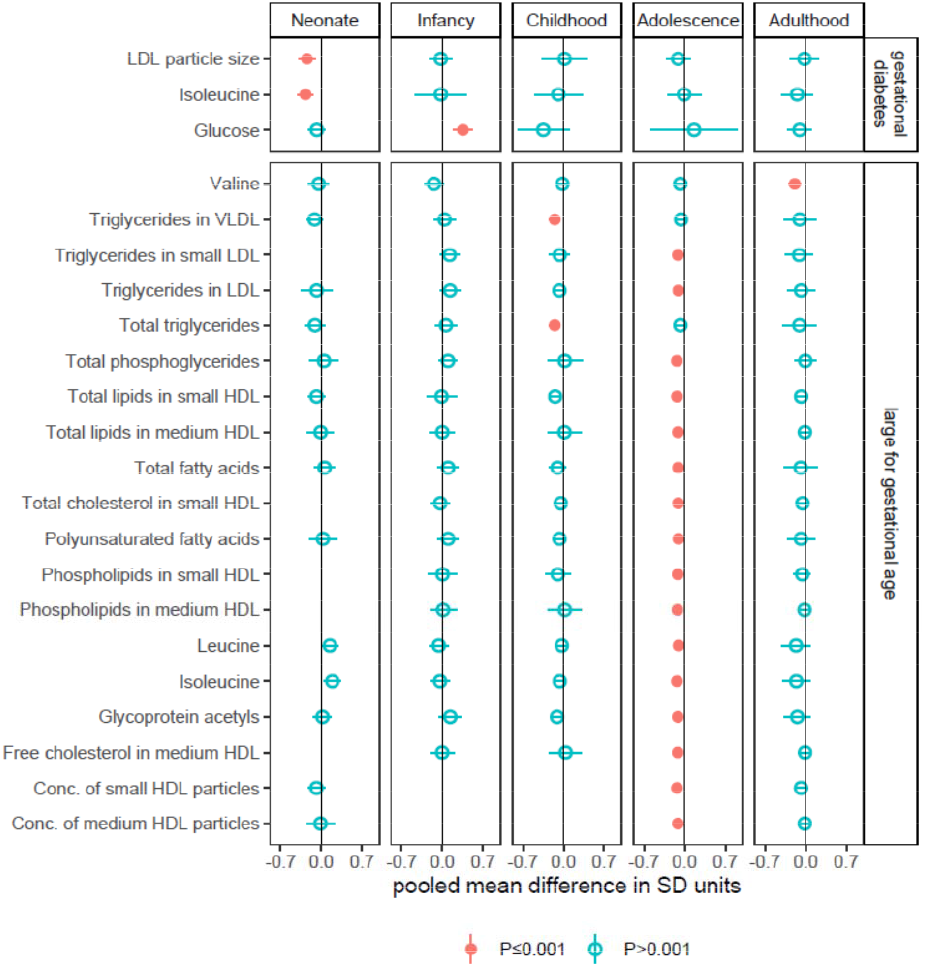
Association of gestational diabetes and large for gestational age with offspring NMR-derived metabolic traits* * Figure shows the pooled adjusted mean differences in standard deviation (SD) units in offspring NMR-derived metabolic traits for gestational diabetes (minus no gestational diabetes), and for large gestational age (minus appropriate size for gestational age), for associations reaching the *P*<0.001 threshold in any one of the five age categories, and equivalent associations in all other age categories (to explore differences by age). Results are adjusted for offspring sex age, and confounders (maternal education, parity age at pregnancy, ethnicity, pre/early pregnancy BMI and smoking). Horizontal bars represent the 95% confidence intervals. Numerical values of these differences are presented in **Table S1**.

Of the nineteen associations of LGA with offspring metabolic traits, none were observed in neonates or infants, two were observed in children, sixteen in adolescents, and one in adults (**Figure 3**). All were inverse associations and represented lower levels of cholesterol, fatty acids, lipoprotein subclasses, three branched-chain amino acids, and the inflammatory marker *glycoprotein acetyls* for LGA (vs. AGA). Associations ranged in magnitude from -0.10SD (95%CI: -0.16 to -0.04, *P*=0.001) for *leucine* in adolescents to -0.19SD (95%CI: -0.29 to - 0.09, *P*=0.0003) for *valine* in adulthood (**Table S1**). For associations seen in adolescence, equivalent associations in adults were slightly attenuated and had wider confidence intervals, and for some traits there also evidence for lower levels in childhood, e.g., difference in *total lipids in small HDL* in childhood, adolescence and adulthood was -0.14SD (95%CI: -0.24 to - 0.04, *P*=0.004), -0.13SD (95%CI: -0.19 to -0.07, *P*=.00006), and -0.08SD (95%CI: -0.16 to 0.01, *P*=0.1), respectively.

Of the 22 associations identified for GD and LGA, only one metabolic trait overlapped with mass spectroscopy measures in the Generation R Study (LGA and child *total triglycerides*). The inverse association in our meta-analysis (difference in child *total triglycerides* for LGA vs. AGA (−0.15SD, 95%CI: -0.24 to -0.06, *P*=0.001) was weaker and imprecisely estimated in Generation R (−0.04SD, 95%CI: -0.40 to 0.32, *P*=0.8, n=339 total with n=75 LGA),

### Overlap in metabolic trait associations

Comparing identified pregnancy/perinatal complication–metabolic trait associations showed that PE/GH were each associated with a different metabolic trait, PTB/SGA were associated with *total HDL cholesterol, total HDL2 cholesterol, and total HDL3 cholesterol* in neonates (higher with PTB and lower with SGA), whereas GD (neonate) and LGA (adolescent) were inversely associated with *isoleucine*. Additionally, SGA (neonates) and LGA (adolescents) were both inversely associated with *concentration of medium HDL particles*, PTB (neonate) and LGA (adolescence) were both inversely associated with *GlycA* and *total lipids in small HDL*, and both PTB (inverse association: neonates) and GD (positive association: infants) were associated with *glucose*

### Between-cohort differences

For most (72%) of the total 3,787 results there was little to no between-cohort heterogeneity (I^2^≤25%, with I^2^=0% for 86% of these), and 8% of the results showed evidence of substantial or high heterogeneity between cohorts (I^2^≥75%) (**Data Set 2**). Four of these were results that met the *P*≤0.001 threshold and all were for PTB neonates (total lipids in small and very large HDL, and concentrations of small and very large HDL particles). Inspecting results from the cohorts contributing to this age (BiB and BIS) uncovered a consistent direction of association in both but mean differences in BiB double those in BIS (e.g., mean difference in *total lipids in small HDL* was -0.86SD in BiB and -0.48SD in BIS. Among other results with substantial or high heterogeneity, 97 were for LGA–adult metabolic traits. Further investigation revealed that this was because LGA was inversely associated with adult biomarkers in ALSPAC (age 24.5 years), with smaller positive associations seen in NFBC1966 (age 46.6 years). Notably, results for NFBC1966 at age 31.2 years were between the ALSPAC age 24.5 years results and the NFBC1966 age 46.6 years results, suggesting a possible age effect (**Figure 4**).

**Figure 4.**
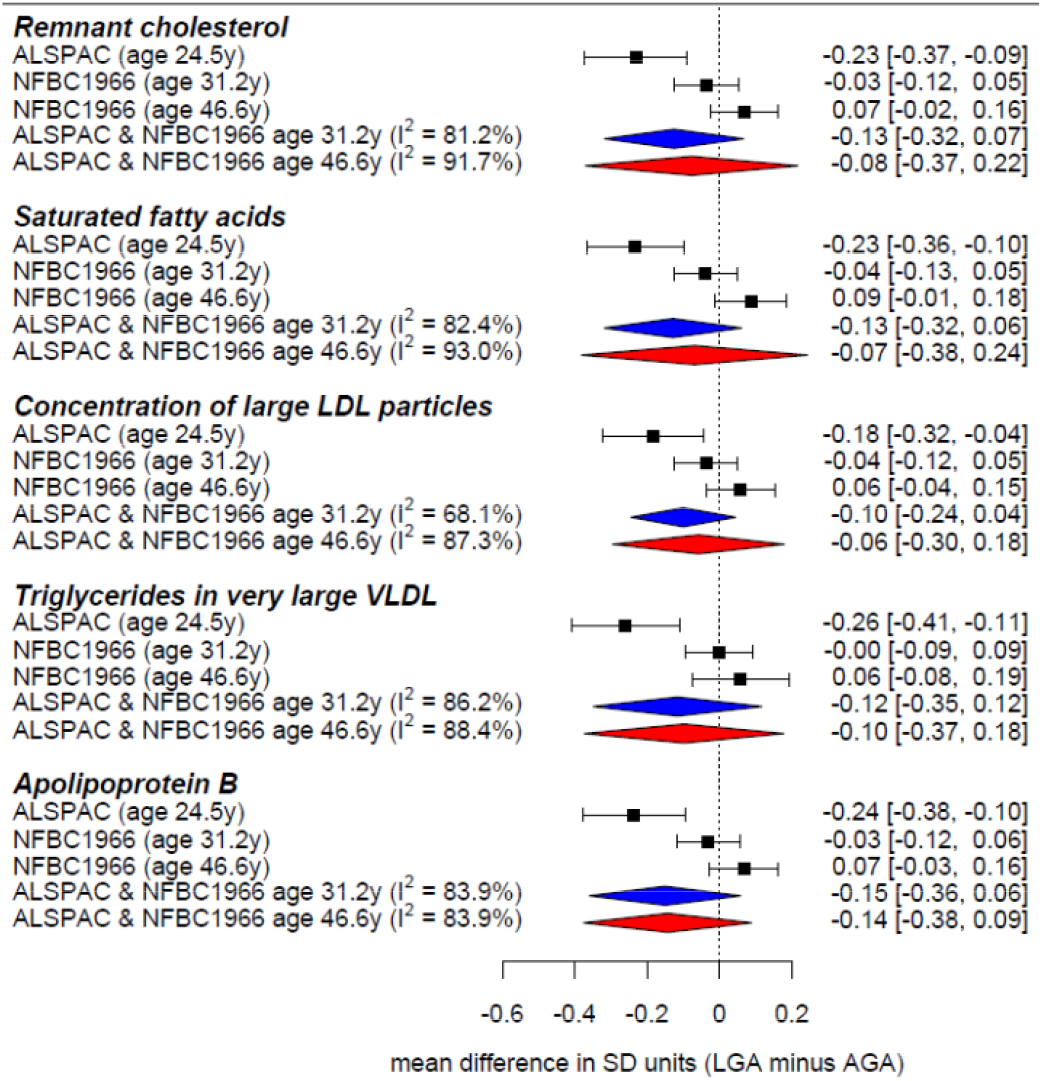
Between-cohort differences in associations of large for gestational age with selected NMR-derived metabolic traits in adults* * Figure shows the cohort-specific and pooled adjusted mean differences in standard deviation (SD) units in five NMR-derived metabolic traits between adults born large gestational age (LGA) and appropriate size for gestational age (AGA). The pooled results from ALSPAC and NFBC19666 (age 46.6y) from the meta-analysis are presented, with the pooled result for ALSPAC and age 31.2y NFBC1966 also presented to highlight differences with age.

### ALSPAC age-change trajectory analysis

Of the twenty associations identified with metabolic traits beyond infancy, one was for SGA, and nineteen were for LGA, and all except one for LGA were included in trajectory analysis from childhood to adulthood in the ALSPAC cohort. Consistent with the meta-analysis, SGA (vs. AGA) was associated with lower *alanine* at baseline though this difference was reduced with increasing child age, with *alanine* slightly higher in SGA from mid adolescence (**Figure S1**). Similarly, most associations between LGA and metabolic traits appeared to change with age from childhood to adulthood (**Figure 5, Figure S2**) e.g., the inverse association observed in the meta-analysis between LGA (vs. AGA) and *triglycerides in VLDL* was attenuated with older age (**Figure 6**).

**Figure 5.**
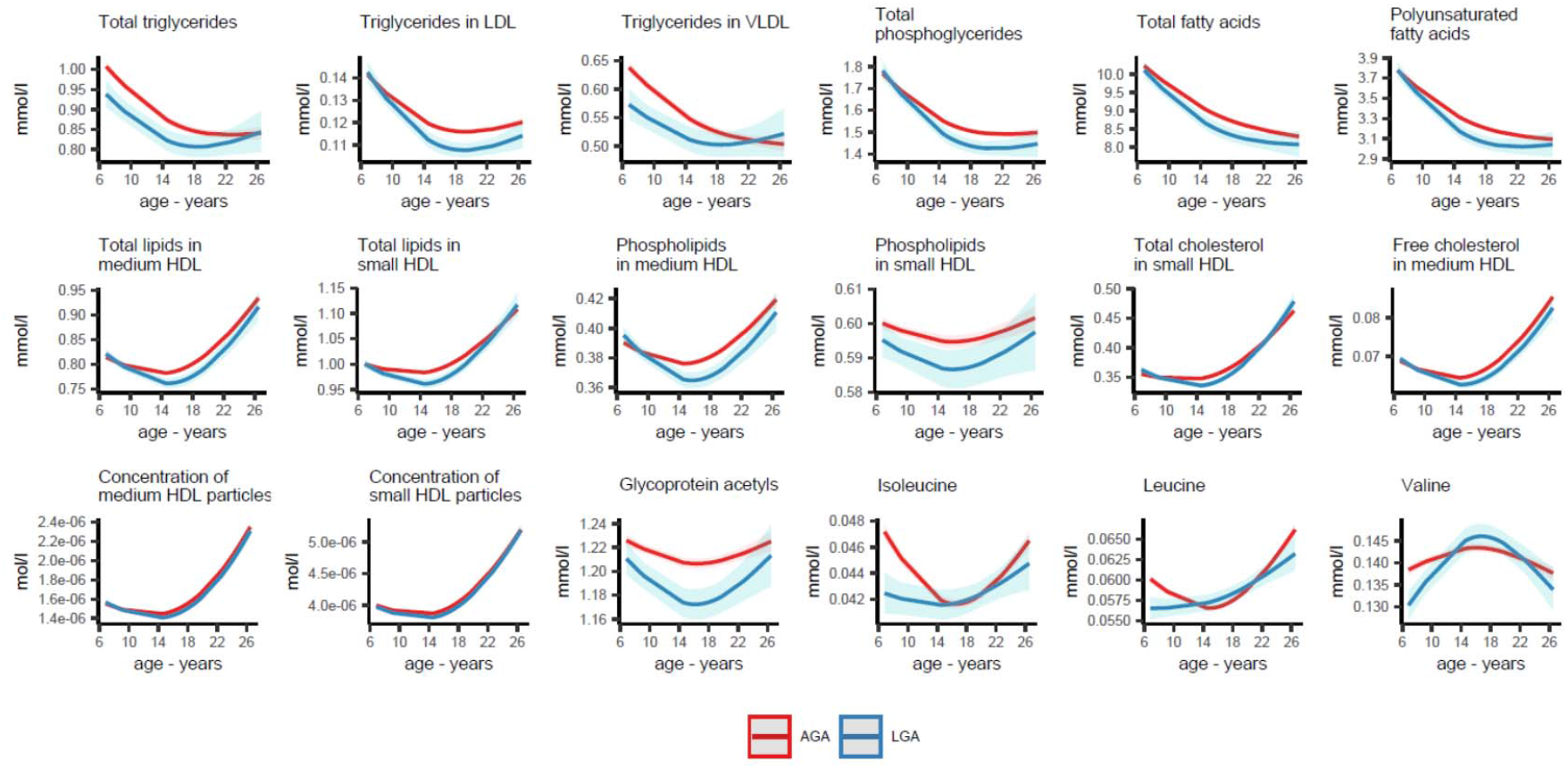
Predicted mean trajectories of NMR-derived metabolic traits from age 7-26 years for offspring born large for gestational age and appropriate size for gestational age from the ALSPAC cohort* * Figure shows the predicted mean NMR-derived metabolic trait trajectories from age 7-26 years for ALSPAC offspring born large for gestational age (LGA, N=500) and appropriate size for gestational age (AGA, N=4,480), for 18 metabolic traits that were identified in the meta-analysis. Predicted values were obtained from adjusted (for sex and confounders) natural cubic spline mixed effects models that included an interaction term with age to allow both LGA/AGA to have different metabolic trait trajectories. Shaded areas represent 95% confidence intervals.

**Figure 6.**
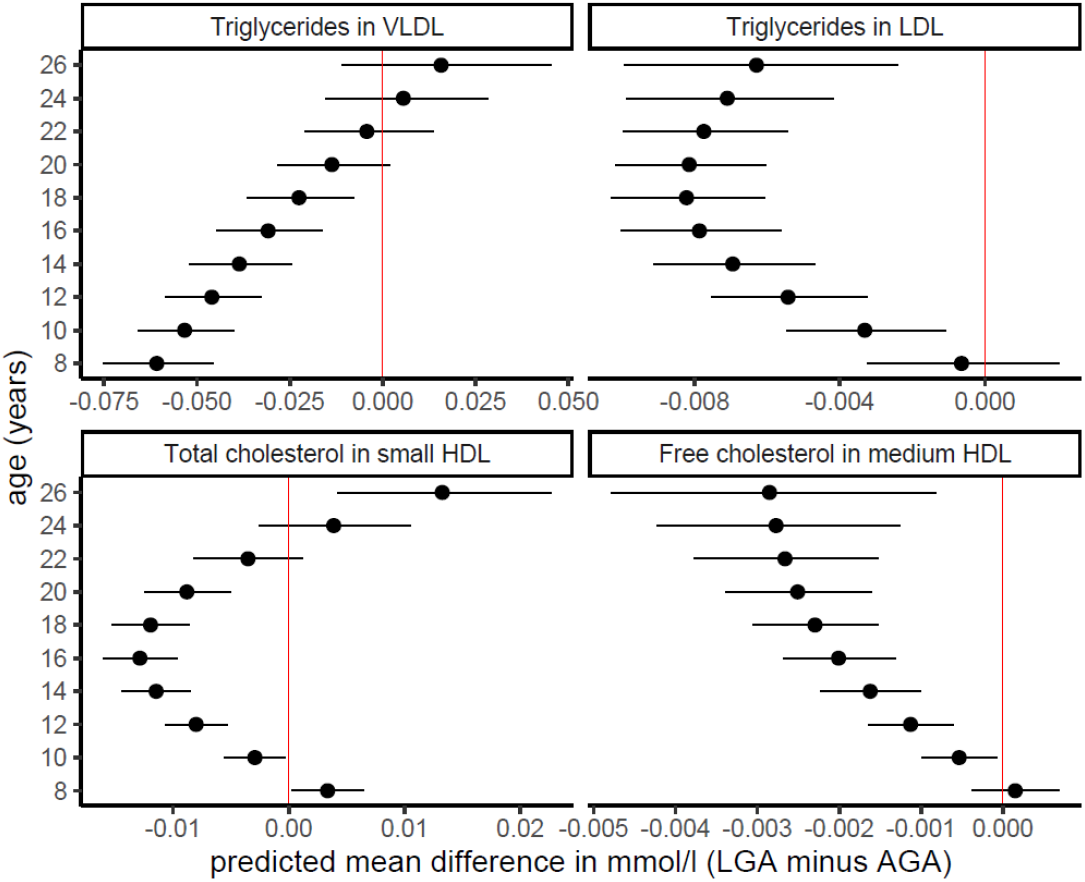
Predicted mean differences in four selected NMR-derived metabolic traits from childhood to adulthood for offspring born large for gestational age and appropriate size for gestational age from the ALSPAC cohort* * Figure shows the predicted mean differences in four NMR-derived metabolic traits between large for gestational age (LGA) and appropriate size for gestational age (AGA) offspring from the ALSPAC cohort. Predicted differences were obtained from adjusted (for sex and confounders) natural cubic spline mixed effects models with an age interaction term to allow LGA/AGA to have different metabolic trait trajectories. Horizontal bars represent the 95% confidence intervals. See **Figure S2** for predicted mean differences for all 18 metabolic traits.

## DISCUSSION

We examined association between common pregnancy/perinatal complications and NMR-derived metabolic profile across the offspring life course in eight population-based cohort studies. Pregnancy complications (PE, GH, GD) were related to only a few metabolic traits mostly in neonates, with little evidence of persistence. More extensive disruptions in neonate metabolic traits were seen for perinatal complications, mostly for PTB, with limited evidence of persistence to older ages, except for differences with LGA which mostly emerged during adolescence and appeared to weaken by older adulthood.

To the best of our knowledge, ours is the first study to simultaneously investigate short- and long-term effects of common pregnancy/perinatal complications on offspring metabolomics. Most associations with metabolic traits were observed in preterm neonates, which appears to support earlier observations of widespread gestational age-dependent effects on metabolites from an untargeted metabolomics analysis in 298 neonates (49) Most pregnancy and perinatal complications were associated with metabolic traits in neonates only with little evidence of persistence beyond early life, This is consistent with Mendelian randomization studies that show that intrauterine exposures related to variation in birth weight are unlikely to causally influence future metabolic health in the offspring (50, 51).

Our finding of inverse associations of LGA with adolescent metabolic traits are consistent with a study of 18,288 adolescents and adults with the same metabolomics platform as our study (52) which showed a higher mean birth weight across the distribution was associated with healthier lipid profile, including lower triglycerides, and other lipids that we see inverse associations of LGA with in our study. This may reflect an interplay with offspring adiposity and puberty whereby LGA offspring have higher prepubertal body fat compared to AGA (53) with this leading to earlier puberty for LGA (54, 55) which in turn might influence metabolic traits (56). This may also explain our finding that the association between LGA and healthier metabolic profile was weakened or even reversed by early midlife (since all AGA offspring should have already completed puberty).

Strengths of this work include the larger sample size compared with previous studies and the examination of metabolic traits during important life periods including whether these change over the life course. Limitations include the small numbers of exposed offspring despite the relatively larger sample size, (e.g., 18.4% exposed to GH – the most common complication). Residual confounding due to use of crude harmonised variables across cohorts (57, 58) and unmeasured confounders (e.g., maternal health) may influence our findings. Complete case analysis may have reduced precision and introduced bias due to missing data, and attrition could introduce a selection bias in the adult results. The *P*-value threshold used to identify robust associations was somewhat arbitrary, so results require replication in other studies. Approaches to select an effective numbers of tests have been developed (59) but to the best of our knowledge have not been extended to multi-cohort analyses, like our study. We have also provided all coefficients and exact *P*-values which readers can use to apply their own *P*-value thresholds (**Data Set 2**). Finally, some effects on metabolic traits may depend on the timing of life course processes such as puberty.

In conclusion, our results offer reassurance that PE, GH, and GD do not result in widespread metabolic disruption in offspring, and that more widespread disruptions for PTB and SGA are mostly confined to neonates. Differences in offspring metabolic traits for LGA require further exploration to establish why they primarily arose during adolescence.

## Data Availability

Data can be accessed by making a formal request to each cohort, see supplement for more details

## ACKNOWLEDGMENTS

We thank all cohort members and researchers who participated in the study. Cohort-specific acknowledgments can be found in **Table S3**.

## SUPPLEMENTAL MATERIAL

### Supplemental Methods

Description of the included cohorts and measurements

**Tables S1** Association of pregnancy and perinatal complications with offspring NMR-derived metabolic traits

**Table S2** Cohort-specific acknowledgements and funding statements

**Figure S1** Predicted mean Alanine trajectories from age 7-26 years and predicted mean differences in ALSPAC offspring born small for gestational age and appropriate size for gestational age

**Figure S2** Predicted mean differences in NMR-derived metabolic traits for born large for gestational age and appropriate size for gestational age offspring from the ALSPAC cohort

**Data Set 1** List of all available offspring NMR-derived metabolic traits in each cohort

**Data Set 2** Complete meta-analysis results i.e., the adjusted pooled mean differences in SD units for all NMR-derived metabolic trait at each age category and by each pregnancy/perinatal complication

## REFERENCES

1. Wang Q, Würtz P, Auro K, et al. Metabolic profiling of pregnancy: cross-sectional and longitudinal evidence. BMC Med. 2016;14(1):205–. doi:10.1186/s12916-016-0733-0

2. Liang L, Rasmussen M-LH, Piening B, et al. Metabolic dynamics and prediction of gestational age and time to delivery in pregnant women. Cell. 2020;181(7):1680–92.e15. doi: 10.1016/j.cell.2020.05.002

3. Kelly RS, Giorgio RT, Chawes BL, et al. Applications of metabolomics in the study and management of preeclampsia; a review of the literature. Metabolomics. 2017;13(7):86. doi:10.1007/s11306-017-1225-8

4. McBride N, Yousefi P, Sovio U, et al. Do mass spectrometry-derived metabolomics improve the prediction of pregnancy-related disorders? Findings from a UK birth cohort with independent validation. Metabolites. 2021;11(8). doi:10.3390/metabo11080530

5. McBride N, Yousefi P, White SL, et al. Do nuclear magnetic resonance (NMR)-based metabolomics improve the prediction of pregnancy-related disorders? Findings from a UK birth cohort with independent validation. BMC Med. 2020;18(1):366. doi:10.1186/s12916-020-01819-z

6. Sovio U, Goulding N, McBride N, et al. A maternal serum metabolite ratio predicts fetal growth restriction at term. Nat Med. 2020;26(3):348–53. doi:10.1038/s41591-020-0804-9

7. Sovio U, McBride N, Wood AM, et al. 4-Hydroxyglutamate is a novel predictor of pre-eclampsia. Int J Epidemiol. 2019;49(1):301–11. doi:10.1093/ije/dyz098

8. Taylor K, Ferreira DLS, West J, Yang T, Caputo M, Lawlor DA. Differences in pregnancy metabolic profiles and their determinants between White European and South Asian women: Findings from the Born in Bradford Cohort. Metabolites. 2019;9(9):190. doi:10.3390/metabo9090190

9. Parikh NI, Gonzalez JM, Anderson CAM, et al. Adverse Pregnancy outcomes and cardiovascular disease risk: unique opportunities for cardiovascular disease prevention in women: a scientific statement from the American Heart Association. Circulation. 2021;143(18):e902–e16. doi:doi:10.1161/CIR.0000000000000961

10. Brosens I, Pijnenborg R, Vercruysse L, Romero R. The “Great Obstetrical Syndromes” are associated with disorders of deep placentation. Am J Obstet Gynecol. 2011;204(3):193–201. doi:10.1016/j.ajog.2010.08.009

11. Kaitu’u-Lino TJ, MacDonald TM, Cannon P, et al. Circulating SPINT1 is a biomarker of pregnancies with poor placental function and fetal growth restriction. Nat Commun. 2020;11(1):2411. doi:10.1038/s41467-020-16346-x

12. Brand JS, West J, Tuffnell D, et al. Gestational diabetes and ultrasound-assessed fetal growth in South Asian and White European women: findings from a prospective pregnancy cohort. BMC Med. 2018;16(1):203. doi:10.1186/s12916-018-1191-7

13. Li M, Hinkle SN, Grantz KL, et al. Glycaemic status during pregnancy and longitudinal measures of fetal growth in a multi-racial US population: a prospective cohort study. Lancet Diabetes Endocrinol. 2020;8(4):292–300. doi:10.1016/s2213-8587(20)30024-3

14. Barker DJ. The origins of the developmental origins theory. J Intern Med. 2007;261(5):412–7. doi:10.1111/j.1365-2796.2007.01809.x

15. Gluckman PD, Hanson MA, Cooper C, Thornburg KL. Effect of in utero and early-life conditions on adult health and disease. N Engl J Med. 2008;359(1):61–73. doi:10.1056/NEJMra0708473

16. Paynter NP, Balasubramanian R, Giulianini F, et al. Metabolic predictors of incident coronary heart disease in women. Circulation. 2018;137(8):841–53. doi:10.1161/CIRCULATIONAHA.117.029468

17. Bavineni M, Wassenaar TM, Agnihotri K, Ussery DW, Lüscher TF, Mehta JL. Mechanisms linking preterm birth to onset of cardiovascular disease later in adulthood. Eur Heart J. 2019;40(14):1107–12. doi:10.1093/eurheartj/ehz025

18. Alsnes IV, Vatten LJ, Fraser A, et al. Hypertension in pregnancy and offspring cardiovascular risk in young adulthood: prospective and sibling studies in the HUNT Study (Nord-Trøndelag Health Study) in Norway. Hypertension. 2017;69(4):591–8. doi:10.1161/hypertensionaha.116.08414

19. Yu Y, Arah OA, Liew Z, et al. Maternal diabetes during pregnancy and early onset of cardiovascular disease in offspring: population based cohort study with 40 years of follow-up. BMJ. 2019;367: l6398. doi:10.1136/bmj.l6398

20. Jaddoe VWV, de Jonge LL, Hofman A, Franco OH, Steegers EAP, Gaillard R. First trimester fetal growth restriction and cardiovascular risk factors in school age children: population based cohort study. BMJ : Br Med J. 2014;348:g14. doi:10.1136/bmj.g14

21. Kurbasic A, Fraser A, Mogren I, et al. Maternal hypertensive disorders of pregnancy and offspring risk of hypertension: a population-based cohort and sibling study. Am J Hypertens. 2019;32(4):331–4. doi:10.1093/ajh/hpy176

22. Geelhoed JJ, Fraser A, Tilling K, et al. Preeclampsia and gestational hypertension are associated with childhood blood pressure independently of family adiposity measures: the Avon Longitudinal Study of Parents and Children. Circulation. 2010;122(12):1192–9. doi:10.1161/circulationaha.110.936674

23. von Elm E, Altman DG, Egger M, Pocock SJ, Gotzsche PC, Vandenbroucke JP. The Strengthening the Reporting of Observational Studies in Epidemiology (STROBE) statement: guidelines for reporting observational studies. Lancet. 2007;370(9596):1453–7. doi:10.1016/s0140-6736(07)61602-x

24. Jaddoe VWV, Felix JF, Andersen AN, et al. The LifeCycle Project-EU Child Cohort Network: a federated analysis infrastructure and harmonized data of more than 250,000 children and parents. Eur J Epidemiol. 2020;35(7):709–24. doi:10.1007/s10654-020-00662-z

25. Soininen P, Kangas AJ, Würtz P, Suna T, Ala-Korpela M. Quantitative Serum Nuclear Magnetic Resonance Metabolomics in Cardiovascular Epidemiology and Genetics. Circulation: Cardiovasc Genet. 2015;8(1):192–206. doi:doi:10.1161/CIRCGENETICS.114.000216

26. Boyd A, Golding J, Macleod J, et al. Cohort Profile: the ‘children of the 90s’--the index offspring of the Avon Longitudinal Study of Parents and Children. Int J Epidemiol. 2013;42(1):111–27. doi:10.1093/ije/dys064

27. Fraser A, Macdonald-Wallis C, Tilling K, et al. Cohort Profile: the Avon Longitudinal Study of Parents and Children: ALSPAC mothers cohort. Int J Epidemiol. 2013;42(1):97–110. doi:10.1093/ije/dys066

28. Northstone K, Lewcock M, Groom A, et al. The Avon Longitudinal Study of Parents and Children (ALSPAC): an update on the enrolled sample of index children in 2019. Wellcome Open Research. 2019;4:51–. doi:10.12688/wellcomeopenres.15132.1

29. Lawlor DA, Lichtenstein P, Långström N. Association of Maternal Diabetes Mellitus in Pregnancy With Offspring Adiposity Into Early Adulthood. Circulation. 2011;123(3):258–65. doi:doi:10.1161/CIRCULATIONAHA.110.980169

30. Bell JA, Bull CJ, Gunter MJ, et al. Early Metabolic Features of Genetic Liability to Type 2 Diabetes: Cohort Study With Repeated Metabolomics Across Early Life. Diabetes Care. 2020:dc192348. doi:10.2337/dc19-2348

31. Wright J, Small N, Raynor P, et al. Cohort Profile: the Born in Bradford multi-ethnic family cohort study. Int J Epidemiol. 2013;42(4):978–91. doi:10.1093/ije/dys112

32. Taylor K, McBride N, J Goulding N, et al. Metabolomics datasets in the Born in Bradford cohort [version 2; peer review: 1 approved, 1 approved with reservations]. Wellcome Open Research. 2021;5(264). doi:10.12688/wellcomeopenres.16341.2

33. Raitakari OT, Juonala M, Rönnemaa T, et al. Cohort profile: the cardiovascular risk in Young Finns Study. Int J Epidemiol. 2008;37(6):1220–6. doi:10.1093/ije/dym225

34. Kaikkonen JE, Würtz P, Suomela E, et al. Metabolic profiling of fatty liver in young and middle-aged adults: Cross-sectional and prospective analyses of the Young Finns Study. Hepatology. 2017;65(2):491–500. doi:10.1002/hep.28899

35. Robinson O, Carter AR, Ala-Korpela M, et al. Metabolic profiles of socio-economic position: a multi-cohort analysis. Int J Epidemiol. 2021;50(3):768–82. doi:10.1093/ije/dyaa188

36. Nordström T, Miettunen J, Auvinen J, et al. Cohort Profile: 46 years of follow-up of the Northern Finland Birth Cohort 1966 (NFBC1966). Int J Epidemiol. 2021. doi:10.1093/ije/dyab109

37. Santos Ferreira DL, Williams DM, Kangas AJ, et al. Association of pre-pregnancy body mass index with offspring metabolic profile: Analyses of 3 European prospective birth cohorts. PLOS Med. 2017;14(8):e1002376. doi:10.1371/journal.pmed.1002376

38. Eriksson JG, Sandboge S, Salonen MK, Kajantie E, Osmond C. Long-term consequences of maternal overweight in pregnancy on offspring later health: findings from the Helsinki Birth Cohort Study. Ann Med. 2014;46(6):434–8. doi:10.3109/07853890.2014.919728

39. Clifford SA, Davies S, Wake M. Child Health CheckPoint: cohort summary and methodology of a physical health and biospecimen module for the Longitudinal Study of Australian Children. BMJ Open. 2019;9(Suppl 3):3–22. doi:10.1136/bmjopen-2017-020261

40. Ellul S, Wake M, Clifford SA, et al. Metabolomics: population epidemiology and concordance in Australian children aged 11-12 years and their parents. BMJ Open. 2019;9(Suppl 3):106–17. doi:10.1136/bmjopen-2017-020900

41. Pinot de Moira A, Haakma S, Strandberg-Larsen K, et al. The EU Child Cohort Network’s core data: establishing a set of findable, accessible, interoperable and re-usable (FAIR) variables. Eur J Epidemiol. 2021. doi:10.1007/s10654-021-00733-9

42. Chappell LC, Cluver CA, Kingdom J, Tong S. Pre-eclampsia. Lancet. doi:10.1016/S0140-6736(20)32335-7

43. Kiserud T, Piaggio G, Carroli G, et al. The World Health Organization Fetal Growth Charts: A Multinational Longitudinal Study of Ultrasound Biometric Measurements and Estimated Fetal Weight. PLoS Med. 2017;14(1):e1002220. doi:10.1371/journal.pmed.1002220

44. Pearce N, Lawlor DA. Causal inference—so much more than statistics. Int J Epidemiol. 2017;45(6):1895–903. doi:10.1093/ije/dyw328

45. Higgins JP, Thompson SG, Deeks JJ, Altman DG. Measuring inconsistency in meta-analyses. Br Med J. 2003;327(7414):557–60. doi:10.1136/bmj.327.7414.557

46. Viechtbauer W. Conducting Meta-Analyses in R with the metafor Package. Journal of Stat Softw. 2010;36(3):1–48. doi:10.18637/jss.v036.i03

47. Elhakeem A, Hughes RA, Tilling K, et al. Using linear and natural cubic splines, SITAR, and latent trajectory models to characterise nonlinear longitudinal growth trajectories in cohort studies. BMC Med Res Methodol. 2022;22(68). doi:10.1186/s12874-022-01542-8

48. Lüdecke. D. ggeffects: Tidy data frames of marginal effects from regression models. J Open Source Softw. 2018;3(772). doi:10.21105/joss.00772

49. Ernst M, Rogers S, Lausten-Thomsen U, et al. Gestational age-dependent development of the neonatal metabolome. Pediatr Res. 2021;89(6):1396–404. doi:10.1038/s41390-020-01149-z

50. Moen GH, Brumpton B, Willer C, et al. Mendelian randomization study of maternal influences on birthweight and future cardiometabolic risk in the HUNT cohort. Nat Commun. 2020;11(1):5404. doi:10.1038/s41467-020-19257-z

51. Bond TA, Richmond RC, Karhunen V, et al. Exploring the causal effect of maternal pregnancy adiposity on offspring adiposity: Mendelian randomization using polygenic risk scores. BMC Med. 2022;.20(34). doi:10.1186/s12916-021-02216-w

52. Würtz P, Wang Q, Niironen M, et al. Metabolic signatures of birthweight in 18 288 adolescents and adults. Int J Epidemiol. 2016;45(5):1539–50. doi:10.1093/ije/dyw255

53. Taal HR, Vd Heijden AJ, Steegers EA, Hofman A, Jaddoe VW. Small and large size for gestational age at birth, infant growth, and childhood overweight. Obesity (Silver Spring). 2013;21(6):1261–8. doi:10.1002/oby.20116

54. Di Giovanni I, Marcovecchio ML, Chiavaroli V, de Giorgis T, Chiarelli F, Mohn A. Being born large for gestational age is associated with earlier pubertal take-off and longer growth duration: a longitudinal study. Acta Paediatr. 2017;106(1):61–6. doi:10.1111/apa.13633

55. Mumby HS, Elks CE, Li S, et al. Mendelian randomisation study of childhood BMI and early menarche. J Obes. 2011;2011:180729–. doi:10.1155/2011/180729

56. Bell JA, Carslake D, Wade KH, et al. Influence of puberty timing on adiposity and cardiometabolic traits: A Mendelian randomisation study. PLoS Med. 2018;15(8):e1002641. doi:10.1371/journal.pmed.1002641

57. Brand JS, Gaillard R, West J, et al. Associations of maternal quitting, reducing, and continuing smoking during pregnancy with longitudinal fetal growth: Findings from Mendelian randomization and parental negative control studies. PLoS Med. 2019;16(11):e1002972. doi:10.1371/journal.pmed.1002972

58. Macdonald-Wallis C, Tilling K, Fraser A, Nelson SM, Lawlor DA. Established preeclampsia risk factors are related to patterns of blood pressure change in normal term pregnancy: findings from the Avon Longitudinal Study of Parents and Children. J Hypertens. 2011;29(9):1703–11. doi:10.1097/HJH.0b013e328349eec6

59. Peluso A, Glen R, Ebbels TMD. Multiple-testing correction in metabolome-wide association studies. BMC Bioinform. 2021;22(1):67. doi:10.1186/s12859-021-03975-2

