## Supplemental file for "Effect of common pregnancy and perinatal complications on offspring metabolic traits across the life course: a multi-cohort study"

### **Supplemental** **Methods** Description of the included cohorts and measurements

1. ***Avon Longitudinal Study of Parents and Children (ALSPAC)***

ALSPAC is a prospective birth cohort study that recruited all pregnant women residing within the catchment area of 3 National Health Service authorities in southwest England with an expected date of delivery between 1991 and 1992. The initial number of pregnancies enrolled is 14,541. Of the initial pregnancies, there was a total of 14,676 fetuses, resulting in 14,062 live births and 13,988 children who were alive at 1 year of age. When the children were approximately 7 years of age, an attempt was made to bolster the initial sample with eligible cases who had failed to join the study originally. The total sample size for analyses using any data collected after the age of seven is 15,454 pregnancies, and 15,589 foetuses. Of these 14,901 were alive at 1 year of age. Detailed data has been collected from offspring and parents by questionnaires, data extraction from medical records, linkage to health records, and dedicated clinic assessments up to the last completed contact in 2018.

ALSPAC contributed to analyses on all 6 pregnancy and perinatal complications. Trained research midwives abstracted data from obstetric medical records. This included any record of a diagnosis of gestational diabetes at any time during the pregnancy in women without existing diabetes at the start of pregnancy, as well as every blood pressure measurement in the medical records along with their corresponding gestational ages at the time of the blood pressure measurement. Gestational age and birthweight were recorded in the delivery room and abstracted from obstetric records and/or birth notifications. ALSPAC was able to adjust for all the pre-specified confounders. For these, maternal ethnicity, education, and parity, and maternal pre-pregnancy weight, height, and smoking, were obtained during pregnancy using questionnaires. Maternal age at pregnancy/birth was obtained from birth records or pregnancy questionnaires. Harmonised exposure and confounder variables were derived from these data, and full details can be found on the European Union’s (EU) Child Cohort Network’s variable catalogue (<https://lifecycle-project.eu/for-scientists/variable-catalogue/>).

Metabolic traits were assayed at 4 timepoints in ALSPAC using high-throughput proton (1H) Nuclear magnetic resonance (NMR) on plasma samples, at mean age 7.5y, 15.4y, 17.8y (152 traits at each age excluding ratios), and 24.5y (146 traits excluding ratios). **Data Set 1** lists all metabolic traits available at each age in the ALSPAC cohort. The samples used were non-fasting at mean age 7.5y, and fasting samples were used at ages 15.4y, 17.8y, and 24.5y. NMR data were available for 5,525 children at age 7.5y, 3,366 at age 15.4y, 3,167 at age 17.8y, and 3,270 at age 24.5y. ALSPAC contributed to meta-analyses of associations with metabolites measured in childhood (mean age 7.5y, N=6,206with complete data), adolescence (mean age 15.4y, N=2,348) and adulthood (mean age 24.5y, N=2,256). ALSPAC was also used for the trajectory analyses of change in metabolic traits with age.

Ethical approval for the study was obtained from the ALSPAC Ethics and Law Committee and the Local Research Ethics Committees. Consent for biological samples was collected in accordance with the Human Tissue Act (2004). Informed consent for the use of data collected via questionnaires and clinics was obtained from participants following the recommendations of the ALSPAC Ethics and Law Committee at the time

2. ***Born in Bradford Study (BiB)***

BiB is a population-based prospective birth cohort including 12,453 women who experienced 13,776 pregnancies between 2007 and 2011. BiB has almost an equal split between White European and South Asian women, all residing in Bradford, UK. Bradford is a city in the North of England with high levels of socioeconomic deprivation, and the cohort was started due to a high prevalence of poor child health in the city. The BiB study website provides more information, including protocols, questionnaires and information on data access data and a full list of all data (<https://borninbradford.nhs.uk/research/documents-data/>). Mothers, and their partners, recruited into the study provided detailed interview questionnaire data, measurements, and biological samples.

BiB contributed to analyses on all 6 pregnancy and perinatal complications. Data on measures of blood pressure and proteinuria were abstracted from medical records. All women booked for delivery in Bradford are offered a 75-g oral glucose tolerance test comprising fasting and 2h post-load samples at around 26–28 weeks gestation. Birthweight was obtained from hospital birth records and in all participants was recorded immediately following birth using SECA digital scales. Duration of gestation was obtained from hospital birth records and was based on the date of the mother’s last menstrual period which was confirmed by a dating ultrasound at around 12 weeks. BiB was able to adjust for all the pre-specified confounders. For these, maternal ethnicity, education, and smoking, were self-reported by the mother at her recruitment questionnaire interview. Age was obtained for all women at pregnancy booking. Maternal BMI was calculated using the height measured at recruitment and weight measured at first antenatal clinic visit (approximately 12 weeks gestation), and it was extracted from medical records. Parity was taken from medical records. Harmonised exposure and confounder variables were derived from these data, and full details can be found on the EU Child Cohort Network’s variable catalogue (<https://lifecycle-project.eu/for-scientists/variable-catalogue/>).

Profiling of circulating lipids, fatty acids, and metabolites was done by the high-throughput Nightingale targeted NMR platform, Metabolic traits were assayed on samples obtained at birth from cord blood samples (n=76 traits excluding ratios) and (non-fasting) at mean age 1.6y (n=149 traits excluding ratios). **Data Set 1** lists the metabolic traits available at each age in the BiB cohort. Cord blood samples were taken whenever possible and immediately processed and stored at -80°C. Samples were taken in a subgroup of offspring in early childhood. Venous cord blood samples were taken at delivery by the attending midwife. Samples were refrigerated at 4°C in tubes until collected by laboratory staff within 12 hours. Samples were then spun, frozen and stored at -80°C. In total, the BiB study collected 9,604 cord blood plasma samples. Infant metabolomics were performed on blood samples that were collected on a subsample of the BiB cohort; those enrolled into the Allergy and Infection Study. Quality control (QC) of the data were undertaken by Nightingale Health prior to returning metabolite concentrations to BiB.

Ethics approval has been obtained for the main platform study and all individual sub studies from the Bradford Research Ethics Committee. Mothers and their partners consented to the linkage of theirs and their child’s data.

3. ***Young Finns Study (YFS)***

YFS comprises a cross-sectional study of 3- to 18-year-old subjects in 1980, and follow-up studies in 1983 and 1986 in conducted in five Finnish university hospitals and in 1989 in one of the study areas (Turku). The study began in 1980, when 3,596 children and adolescents (83.1% of those invited) aged 3, 6, 9, 12, 15, and 18 years randomly chosen from the Finnish national population register participated in the first clinical examinations. When they reached adulthood, follow-up examinations were carried out in 2001, 2007, and 2011. The study has comprised questionnaire data on general health and living conditions, physical activity, eating habits, smoking, psychological variables, and others. Physical examinations covered height, weight, skinfold thickness, pubertal stages and blood pressure. Blood specimens were obtained.

YFS contributed to analyses on 4 pregnancy and perinatal complications (but YFS did not have data on SGA and LGA). YFS was able to adjust for all the pre-specified confounders. Maternal pregnancy smoking was coded as ‘less often than daily or never vs. daily’ in YFS. All 4 variables on pregnancy and perinatal complications and all other confounder variables were harmonised according to the EU Child Cohort Network’s variable catalogue (<https://lifecycle-project.eu/for-scientists/variable-catalogue/>).

The high-throughput Nightingale targeted NMR platform was used on YFS serum samples collected in 2001, 2007, and 2011, corresponding to mean age 22.0y (n=169 traits excluding ratios). **Data Set 1** lists the metabolic traits available in the YFS cohort. All metabolites were measured using a single experimental setup that allows for the simultaneous quantification of routine lipids, lipoprotein subclass distributions, fatty acids, as well as other low–molecular weight metabolites, such as amino acids and glycolysis-related metabolites.

All participants provided written informed consent, and the study was approved by the local ethics committees (Hospital District of Southwest Finland). The study protocol conformed to the ethical guidelines of the 1975 Declaration of Helsinki.

4. ***Northern Finland Birth Cohorts 1986 and 1966 (NFBC1986 and NFBC1966)***

The Northern Finland Birth Cohort studies (<http://www.oulu.fi/nfbc>) are two longitudinal birth cohorts established to study factors affecting preterm birth and consequent morbidity in the two northernmost provinces of Finland, Oulu and Lapland. The NFBC1966 includes 12,058 live births (12,231 children) covering 96% of all eligible births in this region during January-December 1966. Two decades later, a second cohort of 9,432 births (9,479 children) was obtained (NFBC1986) which covered 99% of all the deliveries taking place in the target regions during July 1985-June 1986. In both cohorts, mothers and children have been followed-up since mothers enrolled at their first antenatal clinic visit (10-16th week). For NFBC1986, the 16-year follow-up data collection (2001-2002) included clinical examination and serum collection for 6,621 adolescents (71% of the original cohort). Information on the women was collected by local midwives during routine visits in the free-of-charge communal maternity welfare clinics using a questionnaire. Women visited maternity welfare clinics an average of 7 times during pregnancy, beginning between 10–16 weeks gestation. Maternal questionnaire data was collected at 24 to 28 weeks. Maternal health data was obtained from the antenatal cards filled during routine maternity welfare clinics visits or via questionnaire.

Both NFBC studies contributed to analyses on 5 pregnancy and perinatal complications (but both did not have data on GD). Blood pressure measurements were performed at all maternity welfare clinics visits and documented in the cohort datasets. Both NFBC studies were able to adjust for all the pre-specified confounders, except for ethnicity but the participants were nearly all White Europeans. Maternal height, weight, occupation, smoking status, and parity were collected using questionnaires given to all mothers at their first antenatal clinic visit (16th week of gestation), or in questionnaires administered between the 24th and 28th week of gestation. Harmonised exposure/confounder variables were derived from these data, and details can be found on the EU Child Cohort Network’s variable catalogue (<https://lifecycle-project.eu/for-scientists/variable-catalogue/>).

Participants of both NFBCs fasted overnight before serum collection on the morning of clinic attendance at mean ages 16 in NFBC1986, and 31.2 and 46.6 years in NFBC1966 (n=149 traits at each age excluding ratios). **Data Set 1** lists the metabolic traits available in both NFBC cohorts. All serum samples were stored at -80 °C until thawing and underwent NMR profiling.

Ethical approval for the NFBC1986 and NFBC1966 study was obtained from the Ethics Committee of Northern Ostrobothnia Hospital District, Finland. NFBC1986 received ethical approval from Ethics Committee of Northern Ostrobothnia Hospital District (EETTMK: 108/2017) and Oulu University, Faculty of Medicine, Oulu, Finland.

5. ***Helsinki Birth Cohort Study (HBCS)***

HBCS recruited men and women born at Helsinki University Central Hospital between 1934 and 1944, that attended child welfare clinics in the city and were living in Finland in 1971, when a unique identification number was assigned to all residents of Finland. Most of the children (77%) also went to school in Helsinki.

HBCS contributed to analyses on preterm birth only, with gestational age taken from birth records. HBCS was able to adjust for most pre-specified confounders, except for ethnicity and pregnancy smoking because these were not recorded in HBCS (though almost all participants were white Europeans). Mother’s height and weight prior to delivery were taken from birth records. Father’s education was used as the measure of socioeconomic position because mother’s education not recorded. Preterm birth and remaining confounder variables were harmonised as described in the EU Child Cohort Network’s variable catalogue (<https://lifecycle-project.eu/for-scientists/variable-catalogue/>).

Metabolic traits were assayed at 4 timepoints in HBCS using high-throughput proton (1H) Nuclear magnetic resonance (NMR) on fasting plasma samples, at mean age 66.0y (n=169 traits excluding ratios). **Data Set 1** lists the metabolic traits available in the HBCS cohort.

The clinical study protocol was approved by the Ethics Committee of Epidemiology and Public Health of the Hospital District of Helsinki and Uusimaa. Written informed consent was obtained from each participant before any study procedure was initiated.

6. ***Barwon Infant Study (BIS)***

BIS is a birth cohort study (n = 1064 mother–1074 infant pairs [10 sets of twins]) with antenatal recruitment conducted in the Barwon region in Victoria, Australia. Pregnant women were recruited <28 weeks of gestation between years 2010 and 2013. Detailed questionnaire and clinical data and extensive biospecimens were taken in pregnancy, at birth and at 1, 6, 9 and 12 months, 2- and 4-year.

BIS contributed to analyses on all 6 pregnancy and perinatal complications. Birth weight and gestational age at birth were obtained from hospital records and questionnaires. GD was taken from obtained from hospital records. BIS was able to adjust for all the pre-specified confounders. For these, data on maternal age, maternal smoking during pregnancy were obtained from questionnaires and hospital records. Pre-pregnancy weight was self-reported, and maternal height was measured at the first visit (28–32 weeks’ gestation). Harmonised exposure and confounder variables were derived from these data, and full details can be found on the EU Child Cohort Network’s variable catalogue (<https://lifecycle-project.eu/for-scientists/variable-catalogue/>).

Simultaneous quantification of routine lipids, lipoprotein subclass distributions, particle size and composition, fatty acids, and other low-molecular-weight metabolites such as amino acids and glycolysis-related metabolites was performed using Nightingale high-throughput NMR-based metabolomic platform. Non-fasting samples were obtained at birth from cord blood (n=79 traits excluding ratios), and at ages 1.1 and 4.2 years (n=135 traits excluding ratios at both ages). **Data Set 1** lists the metabolic traits available at each age in the BIS cohort.

Ethics approval was obtained from the Barwon Health Human Research Ethics Committee (10/24). All mothers provided written informed consent

7. ***Longitudinal Study of Australian Children’s Child Health CheckPoint (CheckPoint)***

CheckPoint was conducted between February 2015 and March 2016 at child age of 11–12 years and comprised a detailed cross-sectional assessment of physical health and biomarkers in a population-based national sample of children and their parents from the Growing Up in Australia: Longitudinal Study of Australian Children (LSAC). Child Health CheckPoint was nested between waves 6 (2014) and 7 (2016) of LSAC, a population-based cohort study from early childhood. Of the families agreeing to receive information about the CheckPoint study, 1874 families took part (53% of eligible participants, 42% of Wave 6 cohort and 37% of the original cohort).

CheckPoint contributed to analyses on 5 pregnancy and perinatal complications, except for pre-eclampsia. CheckPoint was able to adjust for all pre-specified confounders except parity and maternal BMI pre/during pregnancy as they were not recorded. Preterm birth, gestational diabetes and hypertension, size for gestational age, maternal smoking and maternal education were obtained from LSAC Wave 1 data. Child age and sex were obtained from Medicare records. Maternal age was self-reported at CheckPoint assessments. Maternal ethnicity was estimated based on SNP genotype data. Harmonised exposure and confounder variables were derived from these data, and full details are found in the EU Child Cohort Network’s variable catalogue (<https://lifecycle-project.eu/for-scientists/variable-catalogue/>.

The Nightingale NMR metabolomics platform was used to obtain metabolomics from the CheckPoint children and parents using the 2016-version quantification algorithm. Metabolites were measured from 0.35 mL of serum using a single high-throughput experimental set up for simultaneous quantification of routine lipids, lipoprotein subclass distributions, particle size and composition, fatty acids, and other low-molecular-weight metabolites such as amino acids and glycolysis-related metabolites. This generated data on 228 metabolite measures in absolute concentration units (e.g., millimoles per litre) and ratios. All metabolic traits were assessed on semi fasted samples (mean 4 hours) at mean age 12.0 years (n=148 traits excluding ratios). **Data Set 1** lists the metabolic traits available in the CheckPoint cohort.

The CheckPoint study was approved by The Royal Children's Hospital Melbourne Human Research Ethics Committee (33225D) and the Australian Institute of Family Studies Ethics Committee (14-26); the latter also provides ethical review and approval for LSAC at every wave. The attending parent/caregiver provided written informed consent for themselves and their child to participate in the study and asked to provide optional consent for the collection and use of biological samples.

8. ***Generation R Study***

The Generation R Study is a population-based prospective cohort study from fetal life until adulthood^34,35^. In total, 9778 mothers with a delivery date from April 2002 until January 2006 were enrolled in the study. Response at baseline was 61%. Extensive assessments were performed in mothers, fathers and their children. In a preselected subsample of the study population, metabolomics analyses were performed. In total, 921 and 503 Dutch children had metabolomics data available at birth (cord blood) and at the age of 10 years, respectively.

Generation R contributed to the replication analyses on all pregnancy and perinatal complications, though replication was only required for PB, SGA, and LGA. Generation R was able to adjust for all pre-specified confounders. Information on maternal age, ethnicity, educational level, parity, and smoking were obtained by questionnaire at enrolment. Maternal height and weight were measured, and BMI was calculated at enrolment.

Umbilical venous cord blood samples were collected directly after birth [median gestational age at birth: 40.3 weeks (95% range 36.6, 42.4)] by a midwife or obstetrician. Child’s nonfasting blood samples were obtained by research nurses at the 10-year follow-up visit to the research center [median age: 9.8 years (95% range 9.1, 10.6)]. A targeted metabolomics approach was adopted to determine serum concentrations (μmol/L) of AA, NEFA, PL and Carn AA were analyzed with 1100 high-performance liquid chromatography (HPLC) system (Agilent, Waldbronn, Germany) coupled to a API2000 tandem mass spectrometer (AB Sciex, Darmstadt, Germany). Carn were measured with a 1200 SL HPLC system (Agilent, Waldbronn, Germany) coupled to a 4000QTRAP tandem mass spectrometer from AB Sciex (Darmstadt, Germany). Quality control and pre‑processing is described previously (50).

The Generation R study has been approved by the Medical Ethical Committee of the Erasmus MC, University Medical Center in Rotterdam (MEC-2012-165-NL40020.078.12). Written informed consent was obtained from the parents or legal representatives of the children. Even with consent of the parents, when the child is not willing to participate actively, no measurements are performed.

### **Ethics approvals**

| Cohort name | Ethic approval description |
| --- | --- |
| ALSPAC | Ethical approval for the study was obtained from the ALSPAC Ethics and Law Committee and the Local Research Ethics Committees. Informed consent for the use of data collected via questionnaires and clinics was obtained from participants following the recommendations of the ALSPAC Ethics and Law Committee at the time. At age 18, study children were sent 'fair processing' materials describing ALSPAC’s intended use of their health and administrative records and were given clear means to consent or object via a written form. Data were not extracted for participants who objected, or who were not sent fair processing materials. Ethical approval for the study was obtained from the ALSPAC Law and Ethics committee and local research ethics committees (NHS Haydock REC: 10/H1010/70). |
| BiB | Ethics approval has been obtained for the main platform study and all individual sub studies from the Bradford Research Ethics Committee. Mothers and their partners consented to the linkage of theirs and their child’s data. |
| *YFS* | All participants provided written informed consent, and the study was approved by the local ethics committees (Hospital District of Southwest Finland). The study protocol conformed to the ethical guidelines of the 1975 Declaration of Helsinki. |
| NFBC1986;  NFBC1966 | Ethical approval for the NFBC1986 and NFBC1966 study was obtained from the Ethics Committee of Northern Ostrobothnia Hospital District, Finland. NFBC1986 received ethical approval from Ethics Committee of Northern Ostrobothnia Hospital District (EETTMK: 108/2017) and Oulu University, Faculty of Medicine, Oulu, Finland. |
| HBCS | The clinical study protocol was approved by the Ethics Committee of Epidemiology and Public Health of the Hospital District of Helsinki and Uusimaa. Written informed consent was obtained from each participant before any study procedure was initiated. |
| BIS | Ethics approval was obtained from the Barwon Health Human Research Ethics Committee (10/24). All mothers provided written informed consent |
| CheckPoint | The CheckPoint study was approved by The Royal Children's Hospital Melbourne Human Research Ethics Committee (33225D) and the Australian Institute of Family Studies Ethics Committee (14-26); the latter also provides ethical review and approval for LSAC at every wave. The attending parent/caregiver provided written informed consent for themselves and their child to participate in the study and asked to provide optional consent for the collection and use of biological samples. |
| Generation R | The Generation R study has been approved by the Medical Ethical Committee of the Erasmus MC, University Medical Center in Rotterdam (MEC-2012-165-NL40020.078.12). Written informed consent was obtained from the parents or legal representatives of the children. Even with consent of the parents, when the child is not willing to participate actively, no measurements are performed. |

### **Tables S1** Association of pregnancy and perinatal complications with offspring NMR-derived metabolic traits

|  | **Neonate** | | **Infancy** | | **Childhood** | | **Adolescence** | | **Adulthood** | |
| --- | --- | --- | --- | --- | --- | --- | --- | --- | --- | --- |
|  | Estimate (95%CI) | *P* | Estimate (95%CI) | *P* | Estimate (95%CI) | *P* | Estimate (95%CI) | *P* | Estimate (95%CI) | *P* |
| ***Pre-eclampsia*** |  |  |  |  |  |  |  |  |  |  |
| Phenylalanine | 0.03 (-0.17 to 0.24) | 0.7 | -0.44 (-0.67 to -0.22) | 0.0002 | -0.04 (-0.40 to 0.32) | 0.8 | -0.17 (-0.34 to -0.01) | 0.03 | -0.06 (-0.24 to 0.12) | 0.5 |
| ***Gestational hypertension*** |  |  |  |  |  |  |  |  |  |  |
| Acetate | 0.03 (-0.10 to 0.16) | 0.7 | -0.43 (-0.59 to -0.28) | 3.7x10^-8^ | -0.19 (-0.62 to 0.24) | 0.4 | -0.05 (-0.17 to 0.08) | 0.5 | 0.0 (-0.08 to 0.08) | 0.9 |
| ***Preterm birth*** |  |  |  |  |  |  |  |  |  |  |
| Glutamine | 0.56 (0.36 to 0.76) | 2.4x10^-8^ | -0.02 (-0.34 to 0.3) | 0.9 | -0.11 (-0.23 to 0.01) | 0.07 | 0.02 (-0.09 to 0.12) | 0.8 | 0.08 (-0.04 to 0.2) | 0.2 |
| Tyrosine | 0.34 (0.19 to 0.49) | 7.0x10^-6^ | -0.02 (-0.37 to 0.34) | 0.9 | -0.05 (-0.18 to 0.08) | 0.4 | 0.01 (-0.13 to 0.16) | 0.9 | -0.02 (-0.12 to 0.07) | 0.6 |
| Total lipids in very small VLDL | 0.31 (0.16 to 0.45) | 0.00004 | -0.06 (-0.34 to 0.22) | 0.7 |  |  | 0.05 (-0.04 to 0.14) | 0.3 | 0.07 (-0.04 to 0.17) | 0.2 |
| Total lipids in small LDL | 0.33 (0.16 to 0.50) | 0.0001 | -0.11 (-0.3 to 0.08) | 0.3 | 0.07 (-0.08 to 0.22) | 0.3 | 0.06 (-0.03 to 0.16) | 0.2 | 0.14 (0.04 to 0.24) | 0.004 |
| Total lipids in small HDL | -0.69 (-1.06 to -0.31) | 0.0003 | -0.06 (-0.21 to 0.08) | 0.4 |  |  | 0.07 (-0.02 to 0.17) | 0.1 | 0.05 (-0.07 to 0.17) | 0.4 |
| Total lipids in large HDL | 0.30 (0.15 to 0.45) | 0.0001 | -0.04 (-0.38 to 0.31) | 0.8 | 0.04 (-0.10 to 0.17) | 0.6 | -0.03 (-0.2 to 0.13) | 0.7 | -0.04 (-0.13 to 0.05) | 0.4 |
| Total lipids in IDL | 0.34 (0.18 to 0.5) | 0.00003 | -0.10 (-0.28 to 0.09) | 0.3 | 0.05 (-0.08 to 0.18) | 0.5 | 0.05 (-0.05 to 0.14) | 0.3 | 0.10 (0.0 to 0.20) | 0.06 |
| Glycoprotein acetyls | -0.46 (-0.62 to -0.3) | 9.0x10^-9^ | -0.06 (-0.47 to 0.34) | 0.8 | 0.01 (-0.11 to 0.14) | 0.8 | 0.10 (-0.02 to 0.22) | 0.1 | -0.01 (-0.11 to 0.09) | 0.8 |
| Total cholesterol | 0.36 (0.21 to 0.52) | 6.7x10^-6^ | -0.09 (-0.27 to 0.09) | 0.3 | 0.05 (-0.08 to 0.18) | 0.4 | 0.05 (-0.04 to 0.15) | 0.3 | 0.11 (0.01 to 0.21) | 0.04 |
| Remnant cholesterol | 0.25 (0.1 to 0.4) | 0.0009 | 0.0 (-0.19 to 0.19) | 0.9 | 0.06 (-0.16 to 0.28) | 0.6 | 0.08 (-0.01 to 0.18) | 0.08 | 0.06 (-0.03 to 0.16) | 0.2 |
| Total cholesterol in LDL | 0.29 (0.13 to 0.46) | 0.0005 | -0.12 (-0.32 to 0.07) | 0.2 | 0.05 (-0.08 to 0.18) | 0.5 | 0.06 (-0.04 to 0.15) | 0.2 | 0.14 (0.04 to 0.24) | 0.006 |
| Total cholesterol in HDL | 0.36 (0.2 to 0.52) | 1.2x10^-5^ | -0.05 (-0.36 to 0.26) | 0.7 | 0.02 (-0.15 to 0.18) | 0.9 | -0.03 (-0.17 to 0.11) | 0.7 | 0.0 (-0.09 to 0.09) | 0.9 |
| Total cholesterol in HDL2 | 0.30 (0.15 to 0.46) | 0.0001 | -0.05 (-0.36 to 0.26) | 0.7 | 0.0 (-0.19 to 0.19) | 0.9 | -0.05 (-0.17 to 0.08) | 0.5 | - | - |
| Total cholesterol in HDL3 | 0.48 (0.31 to 0.66) | 6.0x10^-8^ | -0.05 (-0.31 to 0.22) | 0.7 | 0.06 (-0.08 to 0.2) | 0.4 | 0.05 (-0.08 to 0.18) | 0.5 | - | - |
| Esterified cholesterol | 0.31 (0.15 to 0.47) | 0.0001 | -0.11 (-0.29 to 0.07) | 0.3 | 0.05 (-0.08 to 0.18) | 0.5 | 0.04 (-0.05 to 0.14) | 0.4 | 0.11 (0.01 to 0.21) | 0.03 |
| Free cholesterol | 0.49 (0.33 to 0.65) | 1.1x10^-9^ | -0.11 (-0.29 to 0.08) | 0.3 | 0.06 (-0.07 to 0.19) | 0.4 | 0.07 (-0.02 to 0.16) | 0.1 | 0.09 (-0.01 to 0.19) | 0.06 |
| Total cholines | 0.22 (0.09 to 0.35) | 0.0007 | -0.14 (-0.31 to 0.02) | 0.09 | 0.05 (-0.09 to 0.18) | 0.5 | 0.03 (-0.07 to 0.14) | 0.5 | 0.06 (-0.08 to 0.2) | 0.4 |
| Sphingomyelins | 0.39 (0.23 to 0.54) | 6.8x10^-7^ | -0.14 (-0.33 to 0.05) | 0.1 | 0.05 (-0.08 to 0.18) | 0.4 | 0.02 (-0.07 to 0.11) | 0.7 | 0.10 (0.0 to 0.20) | 0.05 |
| Apolipoprotein B | 0.25 (0.10 to 0.40) | 0.001 | -0.01 (-0.2 to 0.19) | 0.9 | 0.04 (-0.11 to 0.18) | 0.6 | 0.11 (0.01 to 0.2) | 0.02 | 0.08 (-0.02 to 0.18) | 0.1 |
| Estimated degree of unsaturation | -0.75 (-0.93 to -0.57) | 3.3x10^-16^ | -0.09 (-0.26 to 0.09) | 0.3 | -0.12 (-0.41 to 0.16) | 0.4 | -0.05 (-0.17 to 0.08) | 0.5 | 0.08 (-0.1 to 0.27) | 0.4 |
| Saturated fatty acids | 0.38 (0.19 to 0.56) | 7.3x10^-5^ | 0.02 (-0.16 to 0.19) | 0.9 | 0.08 (-0.06 to 0.22) | 0.3 | 0.08 (-0.01 to 0.17) | 0.09 | 0.01 (-0.11 to 0.13) | 0.9 |
| Glucose | -0.31 (-0.46 to -0.15) | 0.0001 | 0.14 (-0.04 to 0.32) | 0.1 | 0.07 (-0.19 to 0.33) | 0.6 | -0.02 (-0.09 to 0.06) | 0.7 | -0.05 (-0.16 to 0.06) | 0.4 |
| Citrate | 0.25 (0.12 to 0.38) | 0.0001 | 0.18 (0.01 to 0.35) | 0.03 | -0.05 (-0.18 to 0.08) | 0.4 | -0.02 (-0.12 to 0.08) | 0.7 | 0.07 (-0.04 to 0.17) | 0.2 |
| Creatinine | -0.32 (-0.45 to -0.18) | 4.5x10^-6^ | -0.1 (-0.26 to 0.07) | 0.2 | -0.27 (-0.83 to 0.28) | 0.3 | 0.07 (-0.02 to 0.17) | 0.1 | 0.06 (-0.03 to 0.14) | 0.2 |
| Albumin | -0.89 (-1.1 to -0.69) | 1.3x10^-17^ | 0.01 (-0.15 to 0.18) | 0.9 | -0.02 (-0.15 to 0.12) | 0.8 | 0.05 (-0.07 to 0.17) | 0.4 | 0.02 (-0.08 to 0.12) | 0.7 |
| Conc. of small HDL particles | -0.69 (-1.06 to -0.32) | 0.0003 | - | - | - | - | 0.07 (-0.03 to 0.17) | 0.1 | 0.05 (-0.07 to 0.16) | 0.4 |
| Conc. of very small VLDL particles | 0.25 (0.10 to 0.40) | 0.0008 | - | - | - | - | 0.07 (-0.02 to 0.16) | 0.1 | 0.07 (-0.04 to 0.17) | 0.2 |
| Conc. of IDL particles | 0.33 (0.17 to 0.49) | 0.00005 | - | - | - | - | 0.06 (-0.04 to 0.15) | 0.2 | 0.10 (0.0 to 0.20) | 0.06 |
| Conc. of small LDL particles | 0.33 (0.16 to 0.49) | 0.0001 | - | - | - | - | 0.07 (-0.02 to 0.17) | 0.1 | 0.13 (0.03 to 0.23) | 0.008 |
| Conc. of large HDL particles | 0.29 (0.14 to 0.44) | 0.0002 | - | - | - | - | -0.03 (-0.19 to 0.12) | 0.7 | -0.04 (-0.13 to 0.05) | 0.3 |
| VLDL particle size | -0.40 (-0.55 to -0.25) | 2.8x10^-7^ | 0.14 (-0.04 to 0.32) | 0.1 | 0.02 (-0.22 to 0.26) | 0.9 | 0.15 (0.04 to 0.25) | 0.006 | -0.05 (-0.15 to 0.05) | 0.3 |
| HDL particle size | 0.76 (0.59 to 0.93) | 1.4x10^-18^ | 0.0 (-0.24 to 0.25) | 0.9 | 0.03 (-0.11 to 0.17) | 0.7 | -0.06 (-0.24 to 0.11) | 0.5 | -0.03 (-0.13 to 0.06) | 0.5 |
| ***Small for gestational age*** |  |  |  |  |  |  |  |  |  |  |
| Total cholesterol in VLDL | 0.29 (0.17 to 0.42) | 4.7x10^-6^ | 0.17 (-0.03 to 0.36) | 0.09 | -0.16 (-0.47 to 0.16) | 0.3 | 0.13 (0.01 to 0.24) | 0.03 | 0.04 (-0.07 to 0.15) | 0.5 |
| Total cholesterol in HDL | -0.37 (-0.52 to -0.21) | 2.4x10^-6^ | -0.07 (-0.25 to 0.11) | 0.5 | 0.05 (-0.11 to 0.2) | 0.5 | 0.0 (-0.11 to 0.11) | 0.9 | -0.03 (-0.21 to 0.15) | 0.8 |
| Total cholesterol in HDL2 | -0.37 (-0.52 to -0.22) | 8.2x10^-7^ | -0.07 (-0.25 to 0.11) | 0.4 | 0.06 (-0.1 to 0.21) | 0.5 | -0.01 (-0.12 to 0.1) | 0.8 | -0.03 (-0.21 to 0.14) | 0.7 |
| Total cholesterol in HDL3 | -0.30 (-0.46 to -0.14) | 0.00001 | -0.05 (-0.23 to 0.14) | 0.6 | 0.02 (-0.13 to 0.18) | 0.8 | 0.05 (-0.05 to 0.16) | 0.3 | 0.02 (-0.16 to 0.19) | 0.9 |
| Total lipids in medium HDL | -0.41 (-0.56 to -0.25) | 2.6x10^-7^ | -0.02 (-0.21 to 0.17) | 0.8 | 0.25 (-0.19 to 0.68) | 0.3 | 0.07 (-0.05 to 0.18) | 0.3 | 0.01 (-0.1 to 0.12) | 0.8 |
| Total lipids in very small VLDL | 0.34 (0.20 to 0.47) | 1.4x10^-6^ | 0.15 (-0.05 to 0.35) | 0.1 | 0.01 (-0.12 to 0.15) | 0.8 | 0.17 (0.06 to 0.29) | 0.003 | 0.02 (-0.15 to 0.18) | 0.8 |
| Conc. of very small VLDL particles | 0.35 (0.21 to 0.48) | 5.2x10^-7^ | - | - | - | - | 0.17 (0.05 to 0.29) | 0.005 | 0.02 (-0.13 to 0.17) | 0.8 |
| Conc. of medium HDL particles | -0.40 (-0.55 to -0.24) | 5.0x10^-7^ | - | - | - | - | 0.07 (-0.05 to 0.19) | 0.3 | 0.01 (-0.1 to 0.12) | 0.8 |
| Alanine | 0.09 (-0.04 to 0.23) | 0.2 | 0.04 (-0.13 to 0.22) | 0.6 | -0.25 (-0.38 to -0.11) | 0.0003 | -0.07 (-0.18 to 0.04) | 0.2 | 0.09 (-0.03 to 0.21) | 0.1 |
| Histidine | -0.21 (-0.33 to -0.08) | 0.001 | 0.03 (-0.13 to 0.2) | 0.7 | -0.14 (-0.26 to -0.02) | 0.02 | 0.02 (-0.1 to 0.13) | 0.8 | 0.01 (-0.11 to 0.12) | 0.9 |
| Apolipoprotein A-I | -0.33 (-0.48 to -0.18) | 0.00002 | -0.02 (-0.19 to 0.16) | 0.8 | 0.02 (-0.14 to 0.17) | 0.8 | 0.04 (-0.07 to 0.16) | 0.4 | -0.02 (-0.2 to 0.16) | 0.8 |
| Omega-3 fatty acids | 0.34 (0.16 to 0.52) | 0.0002 | 0.0 (-0.39 to 0.38) | 0.9 | -0.04 (-0.2 to 0.11) | 0.6 | 0.11 (-0.01 to 0.24) | 0.08 | 0.04 (-0.07 to 0.15) | 0.5 |
| ***Gestational diabetes*** |  |  |  |  |  |  |  |  |  |  |
| LDL particle size | -0.25 (-0.39 to -0.10) | 0.0007 | -0.03 (-0.23 to 0.17) | 0.8 | 0.02 (-0.37 to 0.4) | 0.9 | -0.11 (-0.32 to 0.1) | 0.3 | -0.02 (-0.27 to 0.23) | 0.9 |
| Isoleucine | -0.27 (-0.41 to -0.14) | 0.00008 | -0.03 (-0.47 to 0.42) | 0.9 | -0.09 (-0.5 to 0.32) | 0.7 | -0.01 (-0.29 to 0.28) | 0.9 | -0.14 (-0.42 to 0.13) | 0.3 |
| Glucose | -0.08 (-0.23 to 0.07) | 0.3 | 0.35 (0.18 to 0.52) | 0.00005 | -0.35 (-0.79 to 0.10) | 0.1 | 0.16 (-0.58 to 0.91) | 0.7 | -0.10 (-0.30 to 0.10) | 0.3 |
| ***Large for gestational age*** |  |  |  |  |  |  |  |  |  |  |
| Valine | -0.05 (-0.22 to 0.13) | 0.6 | -0.15 (-0.31 to 0.02) | 0.09 | -0.02 (-0.12 to 0.08) | 0.7 | -0.07 (-0.14 to -0.01) | 0.02 | -0.19 (-0.29 to -0.09) | 0.0003 |
| Conc. of medium HDL particles | -0.01 (-0.25 to 0.22) | 0.9 | - | - | - | - | -0.12 (-0.18 to -0.05) | 0.0003 | -0.01 (-0.11 to 0.08) | 0.8 |
| Conc. of small HDL particles | -0.08 (-0.23 to 0.07) | 0.3 | - | - | - | - | -0.13 (-0.19 to -0.07) | 0.00006 | -0.08 (-0.16 to 0.0) | 0.1 |
| Free cholesterol in medium HDL | - | - | 0.0 (-0.21 to 0.20) | 0.9 | 0.04 (-0.24 to 0.32) | 0.8 | -0.12 (-0.18 to -0.05) | 0.0003 | -0.01 (-0.1 to 0.09) | 0.9 |
| Glycoprotein acetyls | 0.02 (-0.14 to 0.18) | 0.8 | 0.14 (-0.06 to 0.33) | 0.2 | -0.11 (-0.21 to -0.01) | 0.03 | -0.12 (-0.18 to -0.05) | 0.0007 | -0.14 (-0.38 to 0.09) | 0.2 |
| Isoleucine | 0.19 (0.04 to 0.34) | 0.01 | -0.04 (-0.21 to 0.13) | 0.6 | -0.06 (-0.16 to 0.03) | 0.2 | -0.13 (-0.2 to -0.06) | 0.0001 | -0.16 (-0.4 to 0.08) | 0.2 |
| Leucine | 0.15 (0 to 0.29) | 0.05 | -0.06 (-0.23 to 0.11) | 0.5 | -0.03 (-0.13 to 0.07) | 0.6 | -0.10 (-0.16 to -0.04) | 0.001 | -0.17 (-0.41 to 0.08) | 0.2 |
| Phospholipids in medium HDL | - | - | 0.01 (-0.21 to 0.24) | 0.9 | 0.02 (-0.26 to 0.31) | 0.9 | -0.12 (-0.18 to -0.06) | 0.0001 | -0.02 (-0.1 to 0.06) | 0.7 |
| Phospholipids in small HDL | - | - | 0.0 (-0.24 to 0.25) | 0.9 | -0.10 (-0.32 to 0.12) | 0.4 | -0.12 (-0.18 to -0.05) | 0.0003 | -0.06 (-0.2 to 0.09) | 0.4 |
| Polyunsaturated fatty acids | 0.03 (-0.21 to 0.26) | 0.8 | 0.10 (-0.07 to 0.27) | 0.3 | -0.07 (-0.17 to 0.03) | 0.2 | -0.11 (-0.17 to -0.04) | 0.001 | -0.08 (-0.32 to 0.16) | 0.5 |
| Total cholesterol in small HDL | - | - | -0.04 (-0.2 to 0.12) | 0.7 | -0.05 (-0.14 to 0.05) | 0.3 | -0.11 (-0.18 to -0.05) | 0.0008 | -0.05 (-0.13 to 0.02) | 0.2 |
| Total fatty acids | 0.06 (-0.12 to 0.24) | 0.5 | 0.1 (-0.08 to 0.27) | 0.3 | -0.10 (-0.24 to 0.04) | 0.2 | -0.11 (-0.18 to -0.05) | 0.0009 | -0.08 (-0.38 to 0.21) | 0.6 |
| Total lipids in medium HDL | -0.01 (-0.24 to 0.22) | 0.9 | 0.0 (-0.22 to 0.22) | 0.9 | 0.02 (-0.27 to 0.31) | 0.9 | -0.11 (-0.18 to -0.05) | 0.0004 | -0.01 (-0.11 to 0.09) | 0.8 |
| Total lipids in small HDL | -0.08 (-0.23 to 0.07) | 0.3 | -0.01 (-0.27 to 0.24) | 0.9 | -0.14 (-0.24 to -0.04) | 0.004 | -0.13 (-0.19 to -0.07) | 0.00006 | -0.08 (-0.16 to 0.01) | 0.1 |
| Total phosphoglycerides | 0.05 (-0.2 to 0.29) | 0.7 | 0.10 (-0.05 to 0.25) | 0.2 | 0.02 (-0.28 to 0.32) | 0.9 | -0.13 (-0.19 to -0.07) | 0.00004 | -0.01 (-0.19 to 0.18) | 1.0 |
| Total triglycerides | -0.11 (-0.29 to 0.07) | 0.2 | 0.06 (-0.13 to 0.25) | 0.5 | -0.15 (-0.24 to -0.06) | 0.001 | -0.07 (-0.14 to 0) | 0.04 | -0.11 (-0.4 to 0.19) | 0.5 |
| Triglycerides in LDL | -0.08 (-0.36 to 0.2) | 0.6 | 0.13 (-0.04 to 0.3) | 0.1 | -0.07 (-0.17 to 0.03) | 0.2 | -0.11 (-0.17 to -0.04) | 0.001 | -0.08 (-0.31 to 0.15) | 0.5 |
| Triglycerides in small LDL | - | - | 0.12 (-0.05 to 0.3) | 0.2 | -0.06 (-0.24 to 0.11) | 0.5 | -0.11 (-0.18 to -0.05) | 0.0009 | -0.11 (-0.34 to 0.12) | 0.4 |
| Triglycerides in VLDL | -0.12 (-0.25 to 0.02) | 0.09 | 0.04 (-0.15 to 0.22) | 0.7 | -0.15 (-0.24 to -0.06) | 0.001 | -0.06 (-0.13 to 0.01) | 0.1 | -0.10 (-0.38 to 0.18) | 0.5 |

Data shows the adjusted pooled mean differences in SD units (95%CIs) in NMR-derived metabolic traits for associations that reach a threshold of P≤0.001 in one of the age categories as well as the equivalent associations in all other age categories (to explore differences by age). Results are adjusted for offspring sex age, and confounders. The results in this table represent the numerical values for the results shown in Figures 1-3.

### **Table S2** Cohort-specific acknowledgements and funding statements

**1. Avon Longitudinal Study of Parents and Children (ALSPAC)**

We are extremely grateful to all the families who took part in this study, the midwives for their help in recruiting them, and the whole ALSPAC team, which includes interviewers, computer and laboratory technicians, clerical workers, research scientists, volunteers, managers, receptionists and nurses. The UK Medical Research Council and Wellcome (Grant ref: 217065/Z/19/Z) and the University of Bristol provide core support for ALSPAC. A comprehensive list of grant funding is available on the ALSPAC website (<http://www.bristol.ac.uk/alspac/external/documents/grant-acknowledgements.pdf>).

Study data were collected and managed using REDCap electronic data capture tools hosted at the University of Bristol.1 REDCap (Research Electronic Data Capture) is a secure, web-based software platform designed to support data capture for research studies.

**2. Born in Bradford Study (BiB)**

BiB receives core funding from the Wellcome Trust (WT101597MA and 223601/Z/21/Z), a joint grant from the UK Medical and Economic and Social Science Research Councils (MR/N024397/1), British Heart Foundation (CS/16/4/32482), and the National Institute of Health Research under its Applied Research Collaboration for Yorkshire and Humber (NIHR200166) and Clinical Research Network research delivery support. BiB is only possible because of the enthusiasm and commitment of the Children and Parents in BiB. We are grateful to all the participants, teachers, school staff, health professionals and researchers who have made BiB happen

**3. Young Finns Study (YFS)**

The Young Finns Study has been financially supported by the Academy of Finland: grants 322098, 286284, 134309 (Eye), 126925, 121584, 124282, 129378 (Salve), 117787 (Gendi), and 41071 (Skidi); the Social Insurance Institution of Finland; Competitive State Research Financing of the Expert Responsibility area of Kuopio, Tampere and Turku University Hospitals (grant X51001); Juho Vainio Foundation; Paavo Nurmi Foundation; Finnish Foundation for Cardiovascular Research ; Finnish Cultural Foundation; The Sigrid Juselius Foundation; Tampere Tuberculosis Foundation; Emil Aaltonen Foundation; Yrjö Jahnsson Foundation; Signe and Ane Gyllenberg Foundation; Diabetes Research Foundation of Finnish Diabetes Association; This project has received funding from the European Union’s Horizon 2020 research and innovation programme under grant agreements No 848146 (To Aition) and No 755320 (TAXINOMISIS); This project has received funding from the European Research Council (ERC) advanced grants under grant agreement No 742927 (MULTIEPIGEN project); Tampere University Hospital Supporting Foundation and Finnish Society of Clinical Chemistry, and the Cancer Foundation Finland.

**4. Northern Finland Birth Cohorts 1986 and 1966 (NFBC1986 and NFBC1966)**

The authors are very grateful to all the participants who took part in the Northern Finland Birth Cohort 1966 and 1986 studies; to the whole study teams, including research staff and all others involved in data collection and processing; and to those in the oversight and management of the studies. NFBC1986 received funding from EU QLG1-CT-2000-01643 (EUROBLCS) Grant no. E51560, NorFA Grant no. 731, 20056, 30167, USA / NIH 2000 G DF682 Grant no. 50945. NFBC1966 31y follow-up received funding from University of Oulu Grant no. 65354, Oulu University Hospital Grant no. 2/97, 8/97, Ministry of Health and Social Affairs Grant no. 23/251/97, 160/97, 190/97, National Institute for Health and Welfare, Helsinki Grant no. 54121, Regional Institute of Occupational Health, Oulu, Finland Grant no. 50621, 54231. NFBC1966 46y follow-up received funding from University of Oulu Grant no. 24000692, Oulu University Hospital Grant no. 24301140, ERDF European Regional Development Fund Grant no. 539/2010 A31592.

**5. Helsinki Birth Cohort Study (HBCS)**

HBCS was supported by Emil Aaltonen Foundation; Finnish Foundation for Diabetes Research; Foundation for Pediatric Research, Novo Nordisk Foundation; Signe and Ane Gyllenberg Foundation; Sigrid Jusélius Foundation; Samfundet Folkhälsan; Finska Läkaresällskapet; Liv och Hälsa; the Academy of Finland supported (grant no. 129369, 129907, 135072, 129255, and 126775); European Commission within the 7th Framework Programme (DORIAN, grant agreement no. 278603); and European Union Horizon 2020 programme (DYNAHEALTH grant no. 633595).

**6. Barwon Infant Study (BIS)**

We thank the BIS participants for the generous contribution they have made to this project. We also thank current and past staff for their efforts in recruiting and maintaining the cohort and in obtaining and processing the data and biospecimens. The establishment work and infrastructure for the BIS was provided by the Murdoch Children’s Research Institute, Deakin University and Barwon Health. Subsequent funding was secured from the National Health and Medical Research Council of Australia, The Jack Brockhoff Foundation, the Scobie Trust, the Shane O’Brien Memorial Asthma Foundation, the Our Women’s Our Children’s Fund Raising Committee Barwon Health, The Shepherd Foundation, the Rotary Club of Geelong, the Ilhan Food Allergy Foundation, GMHBA Limited and the Percy Baxter Charitable Trust, Perpetual Trustees. In-kind support was provided by the Cotton On Foundation and CreativeForce. Research at Murdoch Children’s Research Institute is supported by the Victorian Government's Operational Infrastructure Support Program. This work was also supported by NHMRC Senior Research Fellowships (1064629 to DB; 1045161 to RS) and NHMRC Investigator Grants to DB (1175744).

**7. Longitudinal Study of Australian Children’s Child Health CheckPoint (CheckPoint)**

This paper uses unit record data from Growing Up in Australia, the Longitudinal Study of Australian Children. The study is conducted in partnership between the Department of Social Services (DSS), the Australian Institute of Family Studies (AIFS) and the Australian Bureau of Statistics (ABS). The CheckPoint work was supported by the National Health and Medical Research Council (NHMRC) of Australia [1041352, 1109355]; the Royal Children’s Hospital Foundation [2014-241]; the Murdoch Children’s Research Institute (MCRI); The University of Melbourne, the National Heart Foundation of Australia [100660]; Financial Markets Foundation for Children [2014-055, 2016-310]; the Victoria Deaf Education Institute; and MBIE Catalyst grant (The New Zealand-Australia Life Course Collaboration on Genes, Environment, Nutrition and Obesity (GENO); UOAX1611; to JOS). This work was also supported by NHMRC Senior Research Fellowships (1046518 to MW; 1064629 to DB; 1045161 to RS) and NHMRC Investigator Grants to DB (1175744). Research at the MCRI is supported by the Victorian Government's Operational Infrastructure Support Program. REDCap (Research Electronic Data Capture) electronic data capture tools were used in this study. More information about this software can be found at: [www.projectredcap.org](http://www.projectredcap.org). The authors thank the LSAC and CheckPoint study participants, staff and students for their contributions.

**8. Generation R Study**

The authors gratefully acknowledge the contribution of participants, research collaborators, general practitioners, hospitals, midwives, and pharmacies in Rotterdam. The general design of the Generation R Study is made possible by financial support from the Erasmus MC, University Medical Center, Rotterdam, Erasmus University Rotterdam, Netherlands Organization for Health Research and Development (ZonMw), Netherlands Organisation for Scientific Research (NWO), Ministry of Health, Welfare and Sport and Ministry of Youth and Families. This project received funding from the European Union's Horizon 2020 research and innovation programme (LIFECYCLE, grant agreement No 733206, 2016, EUCANConnect grant agreement No 824989; ATHLETE, grant agreement No 874583, and under the ERA-NET Cofund action (no 727565), European Joint Programming Initiative “A Healthy Diet for a Healthy Life” (JPI HDHL, EndObesity, ZonMW the Netherlands, (no. 529051026)). VWVJ received a European Research Council Consolidator Grant (ERC-2014-CoG-648916). RG received funding of the Dutch Heart Foundation (grant number 2017T013), the Dutch Diabetes Foundation (grant number 2017.81.002), and the Netherlands Organization for Health Research and Development (NWO, ZonMW, grant number 543003109). The study sponsors had no role in the study design, data analysis, interpretation of data, or writing of this report.

### **Figure S1.** Predicted mean Alanine trajectories from age 7-26 years and predicted mean differences in ALSPAC offspring born small for gestational age and appropriate size for gestational age

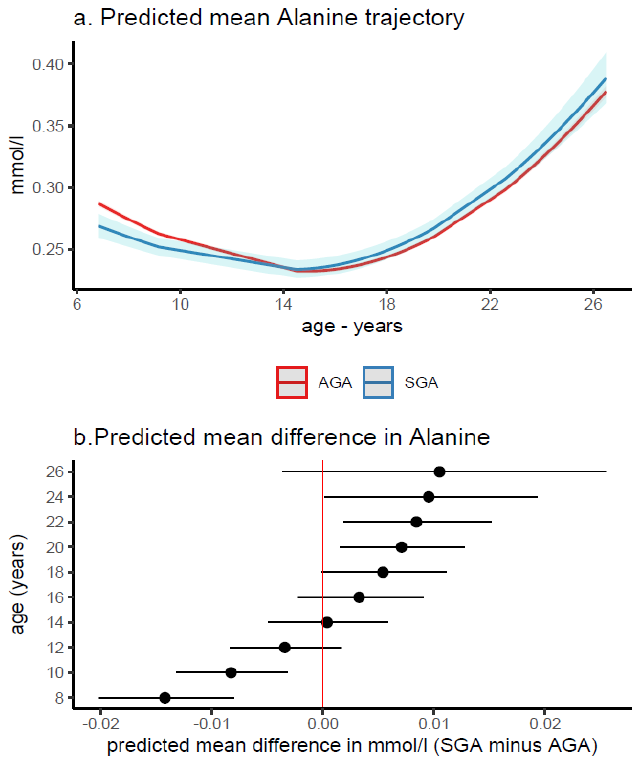

Figure shows (a) predicted mean Alanine trajectory from age 7-26 years and (b) predicted mean difference in ALSPAC offspring born small for gestational age (SGA, N=252) and appropriate size for gestational age (AGA, N=4,480). Predicted values were obtained from adjusted (for sex and confounders) natural cubic spline mixed effects models that included an interaction term with age to allow both SGA/AGA to have different metabolic trait trajectories.

### **Figure S2** Predicted mean differences in NMR-derived metabolic traits for born large for gestational age and appropriate size for gestational age offspring from the ALSPAC cohort

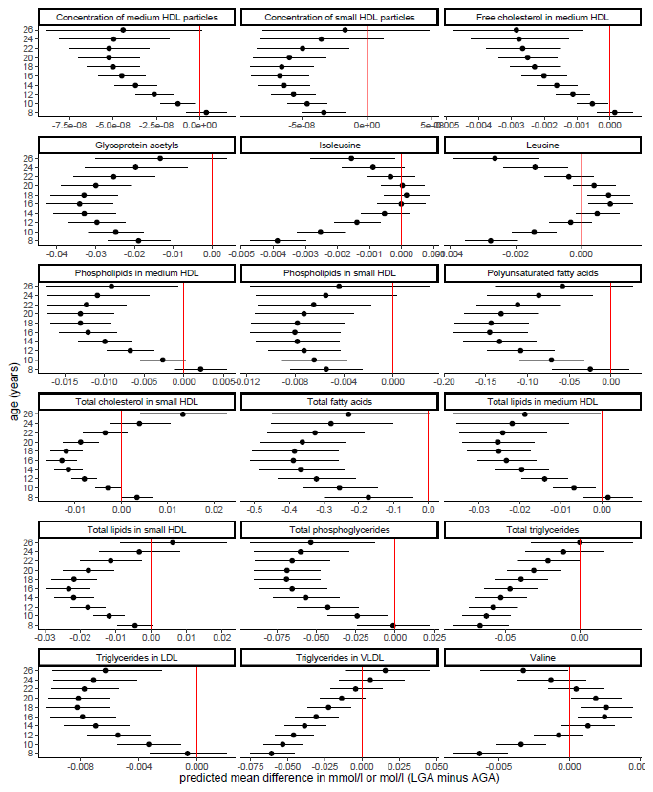

Figure shows the predicted mean differences in NMR-derived metabolic traits between born large for gestational age (LGA, N=500) and appropriate size for gestational age (AGA, N=4,480) offspring from the ALSPAC cohort. Predicted values were obtained from adjusted (for sex and confounders) natural cubic spline mixed effects models that included an interaction term with age to allow both LGA/AGA to have different metabolic trait trajectories.

### **Data Set 1** List of all available offspring NMR-derived metabolic traits in each cohort

See file titled: Data_Set_1_NMR_derived_metabolic_traits_in_each_cohort.csv

### **Data Set 2** Complete meta-analysis results i.e., the adjusted pooled mean differences in SD units for all NMR-derived metabolic trait at each age category and by each pregnancy/perinatal complication

See file titled: Data_Set_2_complete_meta_analysis_results.csv
